# Social functioning in Primary Ciliary Dyskinesia (PCD) – a study of lived experience, relationships and support of patients and caregivers

**DOI:** 10.64898/2026.08.24.26360722

**Authors:** Andrea Fernandez-Rodriguez, Nena Karavasiloglou, Vasiliki Gkatzou, Katie Dexter, Michele Manion, Hansruedi Silberschmidt, Sofia Zambrano, Francesco Pagnini, Living with PCD patient advisory group, Claudia E. Kuehni, Myrofora Goutaki

**Affiliations:** Institute of Social and Preventive Medicine, University of Bern, Bern, Switzerland; Graduate School for Health Sciences (GHS), University of Bern, Bern, Switzerland; PCD Support UK, London, UK; PCD Foundation-Minneapolis, USA; Verein Kartagener Syndrom und Primaere Ciliaere Dyskinesie Deutschland, Wetzikon, Switzerland; Department of Psychology, Università Cattolica del Sacro Cuore, Milan, Italy; Division of Paediatric Respiratory Medicine and Allergology, Department of Paediatrics, Inselspital University Hospital, University of Bern, Bern, Switzerland

**Author notes:** The members of the Living with PCD patient advisory group are listed in the acknowledgements section. **Corresponding author** Myrofora Goutaki, Institute of Social and Preventive Medicine (ISPM), University of Bern, Mittelstrasse 43, 3012 Bern, Switzerland.

**Keywords:** Primary ciliary dyskinesia, rare disease, social participation, lived experience, chronic respiratory disease

## Abstract

Primary ciliary dyskinesia (PCD) is a rare, genetic, multiorgan disease requiring lifelong management. Although PCD affects everyday life, little is known about how people with PCD experience social functioning (SF). We conducted a study within the international participatory *Living with PCD* study to comprehensively explore SF. First, we conducted a focus group and two semi-structured interviews with adults and parents of people with PCD. We analysed qualitative data thematically and used the findings to develop a multilingual online questionnaire on SF.

The questionnaire was completed by 277 participants: 225 adults and adolescents with PCD (81%) and 52 parents of children with PCD (19%). Participants reported active social lives and strong close relationships. PCD had a positive impact on family relationships for 39% of adult/adolescent participants and 41% of parents reporting for children. Among adult/adolescent participants, 49% reported positive or no impact on romantic/intimate relationships, while 17% had avoided or ended a relationship because of PCD. PCD affected the ability to meet responsibilities for 54% of participants, free time for 58%, and planning effort for 53%. Participants were more comfortable discussing PCD with family, friends, and partners than in work or educational settings, where only 29% reported receiving support. Financial support, flexible work, or educational policies and better-trained healthcare professionals were the most frequently identified unmet needs.

This study suggests that maintaining SF with PCD requires substantial individual and relational work. Improving SF for people with PCD requires systemic responses in healthcare, education, and employment, alongside support from close networks.

**Highlights:**

- PCD shapes social functioning beyond health-related quality-of-life.
- Maintaining participation requires planning, adapting activities, and navigating disclosure.
- Close relationships are often supportive, but formal settings pose barriers.
- Work and school support enable disclosure of PCD and more comfortable discussion.
- Structural support is needed to reduce the burden on people with PCD.

## Introduction

Rare disease research has been strongly shaped by efforts to improve diagnosis, clinical management and treatment. However, it largely failed to study the social, geographical, and economic realities that shape the lives of people with rare diseases (1). This broader perspective is particularly relevant for complex chronic diseases that begin early in life and continue into adulthood, where clinical stability does not necessarily translate into unrestricted social functioning (SF).

Primary Ciliary Dyskinesia (PCD) is a rare, genetic, multiorgan disease characterised by impaired ciliary function, which leads to recurrent upper and lower airway infections, progressing to chronic airway impairment and symptoms, needing lifelong demanding treatments (2–4). Over the last 15 years, international collaborations have substantially advanced PCD research, improving diagnosis and understanding PCD’s pathophysiology and natural history of lung disease (5). Yet the everyday social implications of *Living with PCD* remain less well understood. PCD manifestations include chronic cough, bronchiectasis, chronic rhinosinusitis, hearing impairment, laterality defects, and fertility-related issues, and its symptoms may be both visible and misrecognised as common respiratory infection(6–9). This combination of lifelong treatment, broad range of fluctuating symptoms, an increased risk of psychological distress, limited public awareness, and partial visibility makes PCD a particularly relevant case for examining how living with rare chronic diseases shapes SF(10,11).

To date, research on SF in PCD has been scarce and largely limited to its inclusion as a subdomain within broader health-related quality of life (HRQoL) instruments, such as the SF-36, the St. George’s Respiratory Questionnaire (SGRQ) or the disease-specific QoL-PCD (12–15). In the validation of the QoL-PCD instrument, SF had a low mean score, lower than those reported in validation studies of cystic fibrosis (CF) HRQoL instruments (16,17), suggesting that SF may be substantially affected in PCD. However, HRQoL instruments are designed as outcome measures to quantify the presence or severity of problems rather than to explain how these problems are experienced, negotiated, or shaped by context. They therefore provide limited insight into how symptoms, treatment routines, fatigue, infection prevention, fertility concerns, limited disease awareness, and social expectations may shape relationships, education, work, family life, and overall participation in social life.

In our study, we defined SF as the extent to which people with PCD can maintain meaningful relationships, engage in valued social roles, and feel supported and connected in their everyday lives (18,19). SF reflects not only the opportunity and ability to be socially involved but also the personal satisfaction derived from these interactions. Patient support groups have emphasised that comprehensive disease management should include full inclusion, self-determination, and active participation across all life domains (20). People with PCD themselves argue that true health is not simply the absence of illness, but rather the capacity to live a meaningful, socially connected life (21). Yet this broader understanding of SF remains poorly understood in PCD, limiting the evidence available to inform patient-centred care, workplace and educational support, and structural responses. Therefore, we aimed to comprehensively explore how people with PCD and their families experience SF within a large international participatory cohort study. Specifically, we examined relationships, perceived support, management of daily responsibilities, and lived experiences in different settings, and participants’ opinion on structural supports. By doing so, this study seeks to move beyond the measurement of SF as an HRQoL subdomain and to examine how SF in PCD affects social relationships, participation, and support in everyday life.

## Method

### Study design

We conducted a study with two Phases, as this design allowed us to capture lived experience, everyday practices, and social contexts of people with PCD. In Phase 1, we conducted a focus group and two-semi structured interviews with purposively selected people with PCD and parents of people with PCD. Based on the themes identified from these discussions, we developed a cross-sectional quantitative questionnaire (Phase 2). This study design ensured that the questionnaire reflected the SF experience of people with PCD.

We conducted this study within the *Living with PCD*, a participatory international cohort study initiated in 2020 by PCD support groups worldwide and the PCD research team at the University of Bern, Switzerland. The study initially investigated the effects of COVID-19 in PCD (former study name; COVID-PCD; clinicaltrials.gov: NCT04602481) (22). As the cohort expanded from COVID-19-related aims to address broader aspects of living with PCD, it was renamed “*Living with PCD*”. People of any age with either a confirmed or suspected diagnosis of PCD are eligible to participate. Participants register online through the study website (pcd.ispm.ch) and participate anonymously. Adults and adolescents ≥14 years old complete questionnaires themselves; parents provide consent for adolescents where required. For children younger than 14 years, caretakers complete study questionnaires on behalf of their child. In this questionnaire, all caretakers reported that they are parents of the children, so we will use the term parent throughout the manuscript. At registration, participants complete a baseline questionnaire collecting demographic and disease-related information; they are then invited to complete yearly follow-up questionnaires, and additional special thematic questionnaires, including the social functioning questionnaire. The study received ethics approval from the Cantonal Research Ethics Committee in 2020, which was updated in 2025 expanding the study scope (KEK Bern; 2025-00807).

### Focus group discussion and one-to-one interviews

We invited adults diagnosed with PCD or parents of people with PCD to the focus group and one-to-one interviews, aimed to inform the questionnaire development. We used purposive sampling to obtain a diverse set of experiences and invited eight individuals through PCD patient organisations. Six participants attended the focus group. Two were unable to attend the scheduled session so they participated in one-to-one in-depth interviews. All participants provided oral informed consent before starting the discussions.

We developed a semi-structured discussion guide based on a review of the existing literature and iterative discussion within the research team (Supplementary Methods). Questions revolved around 5 key areas of interest: 1) family, friends, intimacy, and support, 2) work, school, and responsibilities, 3) leisure, social life, and belonging, 4) body and self and 5) social differences and inequity. We phrased questions in a way that encouraged reflection on underlying experiences and reasons (“the whys”), while remaining accessible, recognisable, and reflecting everyday life. The focus group discussion took place in June 2025 with AFR and MG as the facilitators and lasted approximately 120 minutes. AFR conducted the individual interviews following the same guide and each lasted 60 minutes. All discussions were guided by our definition of SF (see Introduction), while remining open to additional topics, examples, and meanings that participants considered relevant to SF.

We recorded all sessions with consent and we transcribed the recordings verbatim and imported them into MAXQDA version 24.10 for analysis (23). AFR followed an inductive thematic analysis approach, generating themes directly from the data rather than being based on a pre-existing codebook (24).

In total, eight people participated in the focus group discussion and the one-to-one interviews; 4 of them were people with PCD (three women and one man). The remaining 4 were parents of people with PCD, whose ages ranged from 10 to 35 years. All participating parents were mothers. The age of participants in the discussion ranged between 34 and 66 years old. Using an inductive thematic analysis approach, we developed 4 themes (Figure 1): 1) Having a PCD diagnosis shapes social life, 2) Everyday management and loss of spontaneity, 3) Support and relationship dynamics, and 4) System structures and inequities, which we describe in detail in the Supplementary Results.

**Figure 1.**
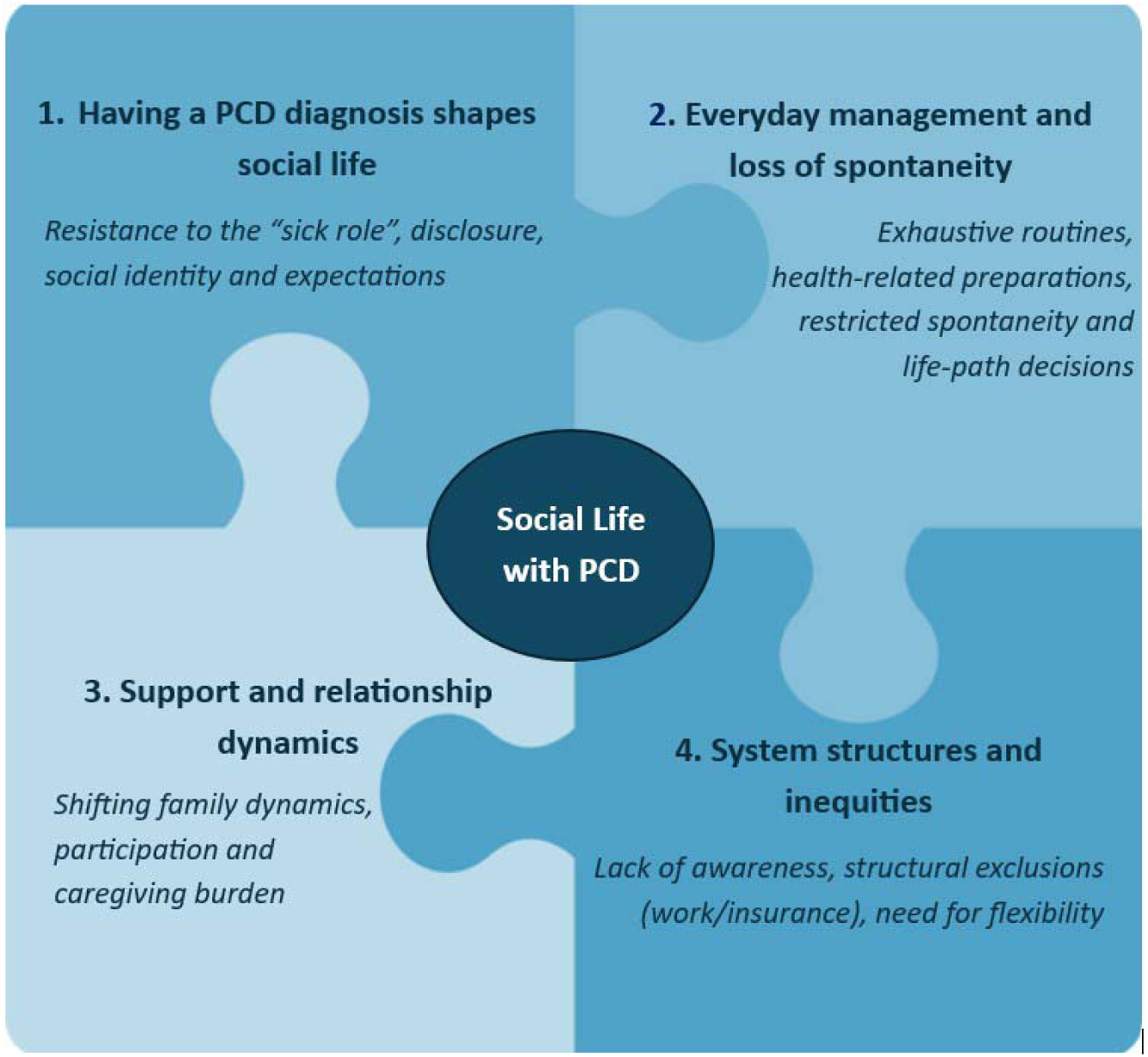
Overview of Social Functioning Themes developed from focus group discussion and one-to-one interviews with people with primary ciliary dyskinesia (PCD) and parents of people with PCD

### Questionnaire development and distribution

Themes from the qualitative analysis directly informed the content of the SF questionnaire we developed. For the questionnaire, we decided to follow the same structure as in the discussion guide as we agreed on it being more intuitive for participants. The questionnaire included 6 sections in which we asked multiple choice and open-ended questions: 1) Basic information, 2) Relationships, 3) Work, school and responsibilities, 4) Free time, 5) How others see you and how you see yourself, and 6) Everyday challenges and support. Participants could skip questions or whole sections of the questionnaire. We first developed the adult version of the questionnaire and then we adapted it for adolescents and parents of children with PCD. The full questionnaire for the adults in English is appended in the Supplementary Methods, for reference. The three age-specific versions of the questionnaire underwent several rounds of revision involving the PCD research team at the University of Bern, external advisors (FP, SZ) and representatives from the *Living with PCD* patient advisory board (KD, MM, HS). The questionnaire was first developed in English and then translated into the other four study languages (German, French, Italian, and Spanish). Native speaking members from patient support groups or our research team performed the translations; two research team members independently checked translations to ensure the meaning stayed the same across languages.

We entered each age- and language-specific version of the questionnaire on the Research Electronic Data Capture (REDCap) platform hosted at the Swiss medical registries and data linkage centre (SwissRDL) at the University of Bern (25). We circulated the SF questionnaire by email on 28th November 2025, to all participants enrolled in the *Living with PCD* study. We additionally advertised the questionnaire via *Living with PCD* study partner patient support organisations. We sent up to three reminders to registered *Living with PCD* study participants who had not completed the questionnaire. We collected responses until 28^th^ February 2026.

### Statistical analysis

We described characteristics of participants using counts and proportions or median and interquartile range (IQR), as appropriate. For the present analysis, year of birth, sex, age at diagnosis and situs abnormalities were obtained from the baseline questionnaire; age at survey was calculated from year of birth and the date of completion of the social functioning questionnaire. Sex was self-reported in response to the question “What is your sex?”, with the response options “male”, “female” and “other”. We did not collect gender identity separately. We also descriptively presented SF aspects separately in adults and adolescents (i.e., those who filled out the questionnaire by themselves) and in parents of children with PCD, and, when possible, overall.

We explored the association between communication behaviours, specifically 1) feeling comfortable discussing about PCD in work or school settings and 2) behaviours related to hiding or justifying symptoms in public, with potentially relevant factors using multivariable logistic regression. We selected factors based on availability and their relevance to disclosure, symptom management in public, and perceived support. The factors we included were age (per 5-year increase), sex, reported support at work or school, and perceived family support. Due to small numbers (n=1), participants selecting “other” in the sex variable were not analysed as a separate category in regression models. We dichotomised both support-related variables as “supported” if participants reported feeling very or somewhat supported, and as “not supported” if they reported feeling neutral, not really supported or not supported at all. All analyses were done with R software (version 4.5.3, for Windows, R Foundation for Statistical Computing).

## Results

### Participants characteristics

Out of the 678 *Living with PCD* study participants with a valid e-mail address to whom we could send the SF questionnaire, 277 submitted a completed questionnaire (41% response rate) (Figure S1). Median age at questionnaire completion was 38 years (IQR 22-54); 64% of respondents were women. Most respondents were adults or adolescents (n=225, 81%); 52 were parents who responded on behalf of their children.

Among the 52 parents responding to the questionnaire, 49 were mothers (94%) and only 3 were fathers. 35 (67%) of responding parents were the main caregiver, i.e., had >50% of caregiving responsibilities. Caregivers shared responsibilities equally in the remaining 33% of cases (17). Information on age at diagnosis, situs abnormalities, country of residence, migrant background, language barrier, current occupation and parental responsibilities are summarised in Table 1.

**Table 1.**
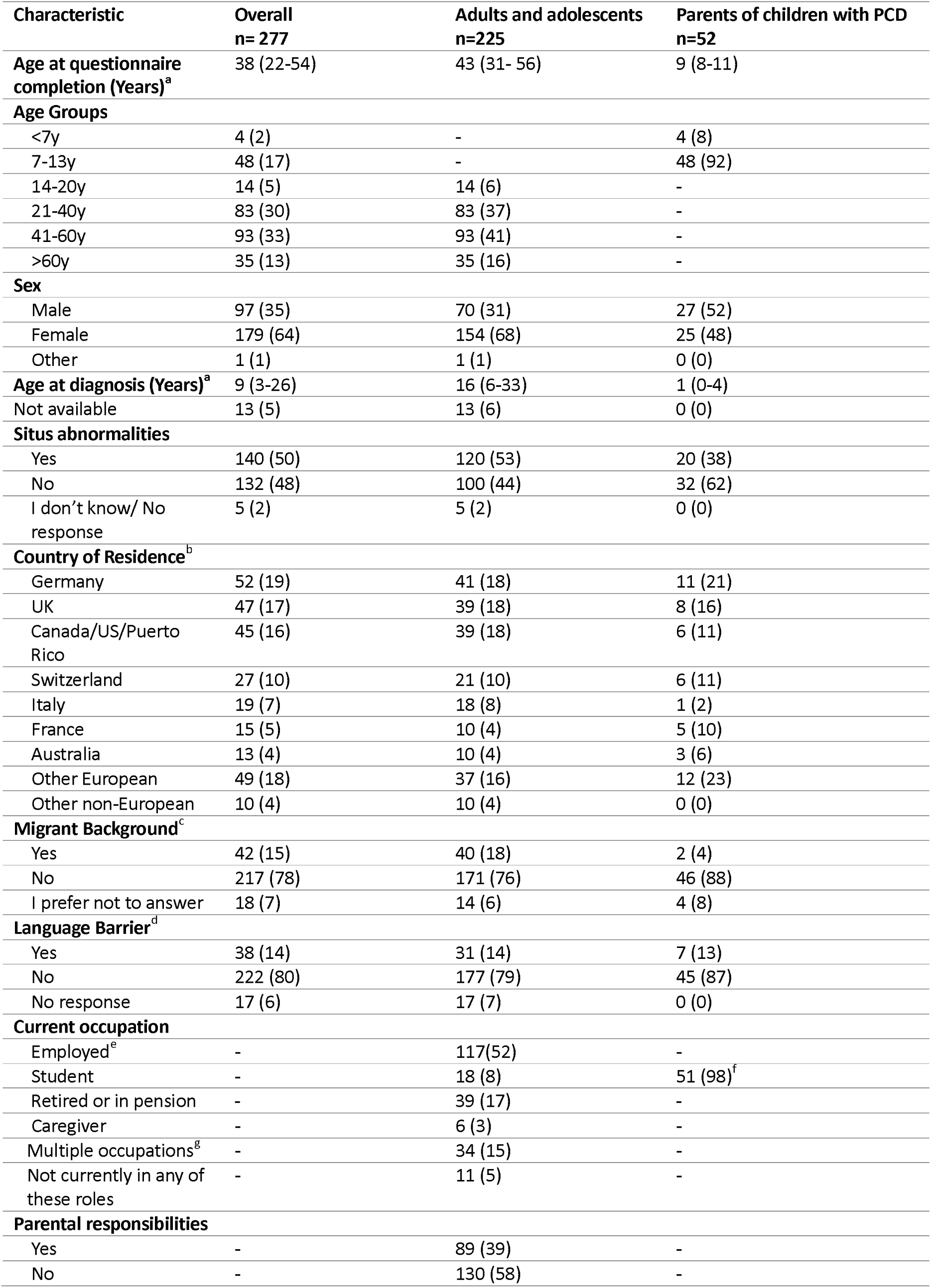
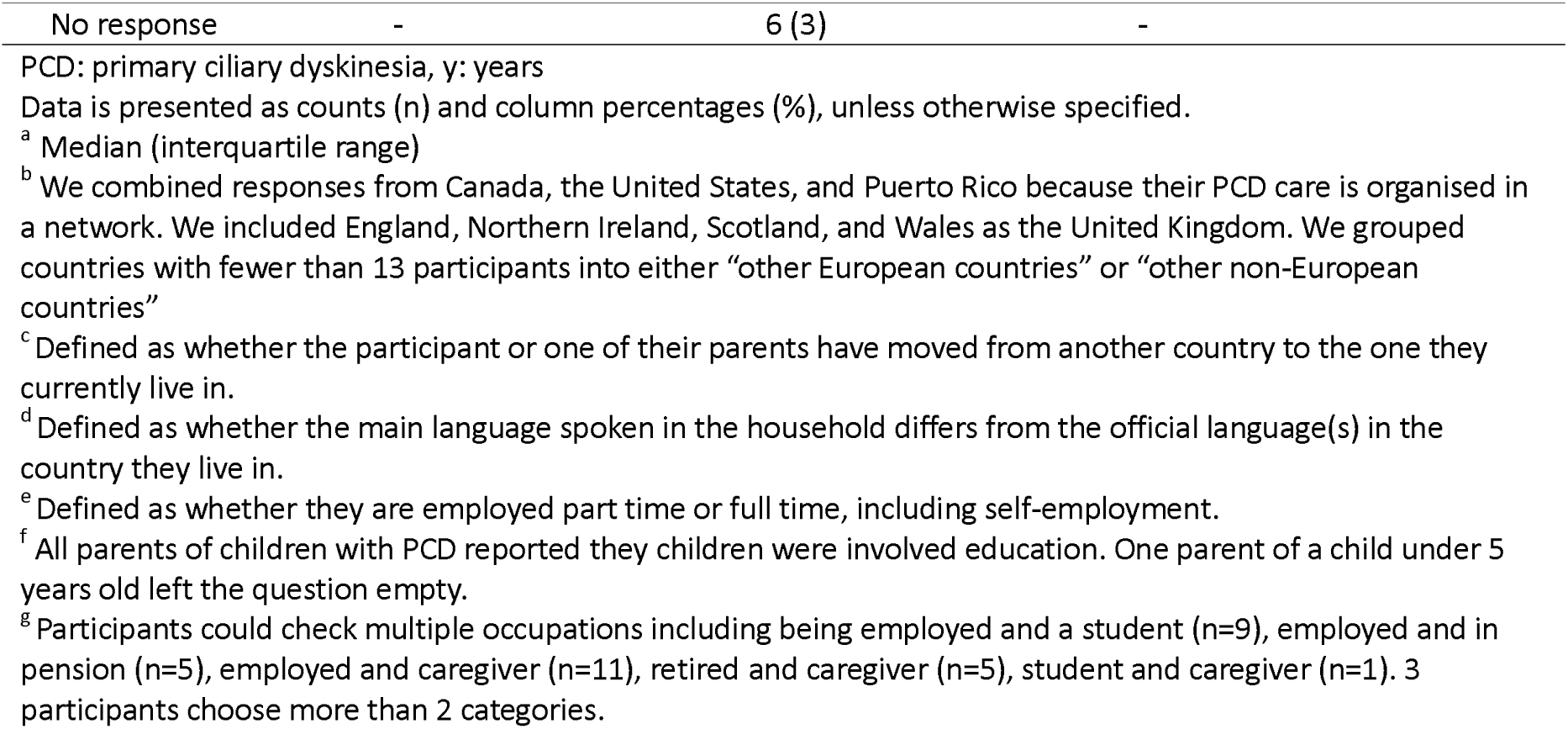
Characteristics of people with primary ciliary dyskinesia registered in the Living with PCD study, who completed the social functioning questionnaire, overall and by age group.

| Characteristic | Overall<br>n= 277 | Adults and adolescents<br>n=225 | Parents of children with PCD<br>n=52 |
| --- | --- | --- | --- |
| **Age at questionnaire completion (Years)<sup>a</sup>** | 38 (22-54) | 43 (31- 56) | 9 (8-11) |
| **Age Groups** |  |  |  |
| <7y | 4 (2) | - | 4 (8) |
| 7-13y | 48 (17) | - | 48 (92) |
| 14-20y | 14 (5) | 14 (6) | - |
| 21-40y | 83 (30) | 83 (37) | - |
| 41-60y | 93 (33) | 93 (41) | - |
| >60y | 35 (13) | 35 (16) | - |
| **Sex** |  |  |  |
| Male | 97 (35) | 70 (31) | 27 (52) |
| Female | 179 (64) | 154 (68) | 25 (48) |
| Other | 1 (1) | 1 (1) | 0 (0) |
| **Age at diagnosis (Years)<sup>a</sup>** | 9 (3-26) | 16 (6-33) | 1 (0-4) |
| Not available | 13 (5) | 13 (6) | 0 (0) |
| **Situs abnormalities** |  |  |  |
| Yes | 140 (50) | 120 (53) | 20 (38) |
| No | 132 (48) | 100 (44) | 32 (62) |
| I don't know/ No response | 5 (2) | 5 (2) | 0 (0) |
| **Country of Residence<sup>b</sup>** |  |  |  |
| Germany | 52 (19) | 41 (18) | 11 (21) |
| UK | 47 (17) | 39 (18) | 8 (16) |
| Canada/US/Puerto Rico | 45 (16) | 39 (18) | 6 (11) |
| Switzerland | 27 (10) | 21 (10) | 6 (11) |
| Italy | 19 (7) | 18 (8) | 1 (2) |
| France | 15 (5) | 10 (4) | 5 (10) |
| Australia | 13 (4) | 10 (4) | 3 (6) |
| Other European | 49 (18) | 37 (16) | 12 (23) |
| Other non-European | 10 (4) | 10 (4) | 0 (0) |
| **Migrant Background<sup>c</sup>** |  |  |  |
| Yes | 42 (15) | 40 (18) | 2 (4) |
| No | 217 (78) | 171 (76) | 46 (88) |
| I prefer not to answer | 18 (7) | 14 (6) | 4 (8) |
| **Language Barrier<sup>d</sup>** |  |  |  |
| Yes | 38 (14) | 31 (14) | 7 (13) |
| No | 222 (80) | 177 (79) | 45 (87) |
| No response | 17 (6) | 17 (7) | 0 (0) |
| **Current occupation** |  |  |  |
| Employed<sup>e</sup> | - | 117(52) | - |
| Student | - | 18 (8) | 51 (98)<sup>f</sup> |
| Retired or in pension | - | 39 (17) | - |
| Caregiver | - | 6 (3) | - |
| Multiple occupations<sup>g</sup> | - | 34 (15) | - |
| Not currently in any of these roles | - | 11 (5) | - |
| **Parental responsibilities** |  |  |  |
| Yes | - | 89 (39) | - |
| No | - | 130 (58) | - |
| No response | - | 6 (3) | - |

PCD: primary ciliary dyskinesia, y: years
Data is presented as counts (n) and column percentages (%), unless otherwise specified.
<sup>a</sup> Median (interquartile range)
<sup>b</sup> We combined responses from Canada, the United States, and Puerto Rico because their PCD care is organised in a network. We included England, Northern Ireland, Scotland, and Wales as the United Kingdom. We grouped countries with fewer than 13 participants into either “other European countries” or “other non-European countries”
<sup>c</sup> Defined as whether the participant or one of their parents have moved from another country to the one they currently live in.
<sup>d</sup> Defined as whether the main language spoken in the household differs from the official language(s) in the country they live in.
<sup>e</sup> Defined as whether they are employed part time or full time, including self-employment.
<sup>f</sup> All parents of children with PCD reported they children were involved education. One parent of a child under 5 years old left the question empty.
<sup>g</sup> Participants could check multiple occupations including being employed and a student (n=9), employed and in pension (n=5), employed and caregiver (n=11), retired and caregiver (n=5), student and caregiver (n=1). 3 participants choose more than 2 categories.

### Impact of PCD on relationships

Most participants reported that PCD either made family relationships stronger (111, 40%) or that it played no significant role (125, 45%, Table 2). In contrast, romantic and intimate relationships were more often negatively impacted (parents were asked for own relationship with the other parent). While 12% (26) of adults/adolescents and 23% (12) of parents felt PCD strengthened their romantic bonds, 38% (84) of adults/adolescents and 42% (22) of parents reported that PCD made romantic relationships “more complicated”. Regarding emotional closeness, 28% (64) of adults/adolescents noted that PCD positively impacted emotional closeness, while 50% (112) reported no impact. Among 189 participants who were in romantic relationships, 20% (37) reported ending or avoiding a relationship due to PCD. The reasoning behind this decision revolved around four main concerns: the fear of being a burden to a partner, fertility-related challenges, lack of support from the partner, and negative self-perception, such as feeling unattractive due to symptoms. Selected participants’ comments regarding these decisions are provided in Textbox S1.

**Table 2.**
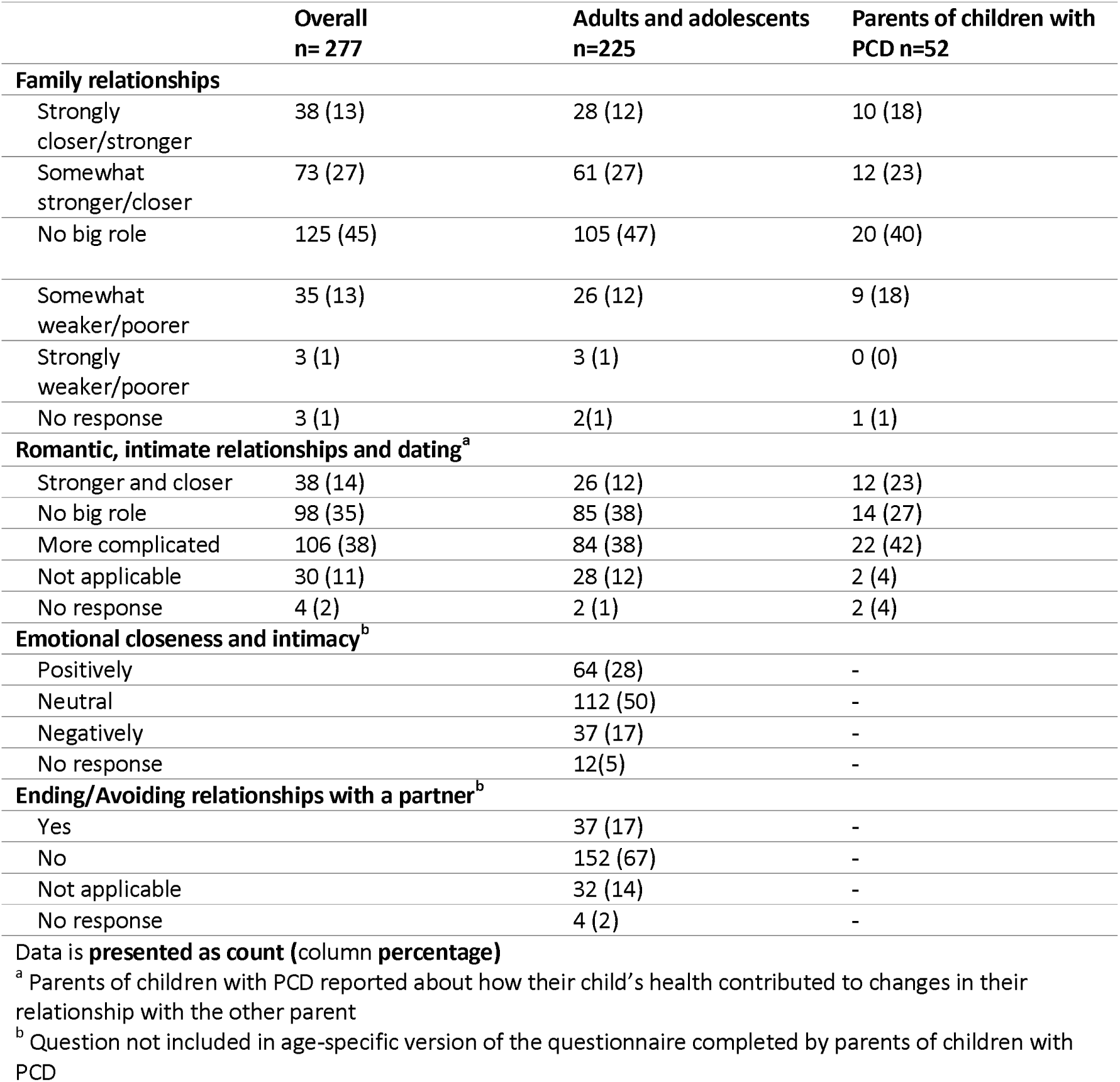
Impact of PCD on the relationships overall and by age group. Responses from people with primary ciliary dyskinesia registered in the Living with PCD study who completed the social functioning questionnaire.

Participants reported how their family members and friends responded to their symptoms and PCD diagnosis (Figure 2). Overall, the most commonly reported responses of family members were “trying to protect the person with PCD or doing things for them” (127, 46%), followed by getting “anxious/stressed when they (i.e., the person with PCD) showed symptoms around them (family members)” (108, 39%). Regarding friendship dynamics, around one third of participants reported friends being supportive and checking in on the person with PCD, considering their limitations when planning activities, and asking questions and try to learn more about PCD.

**Figure 2.**
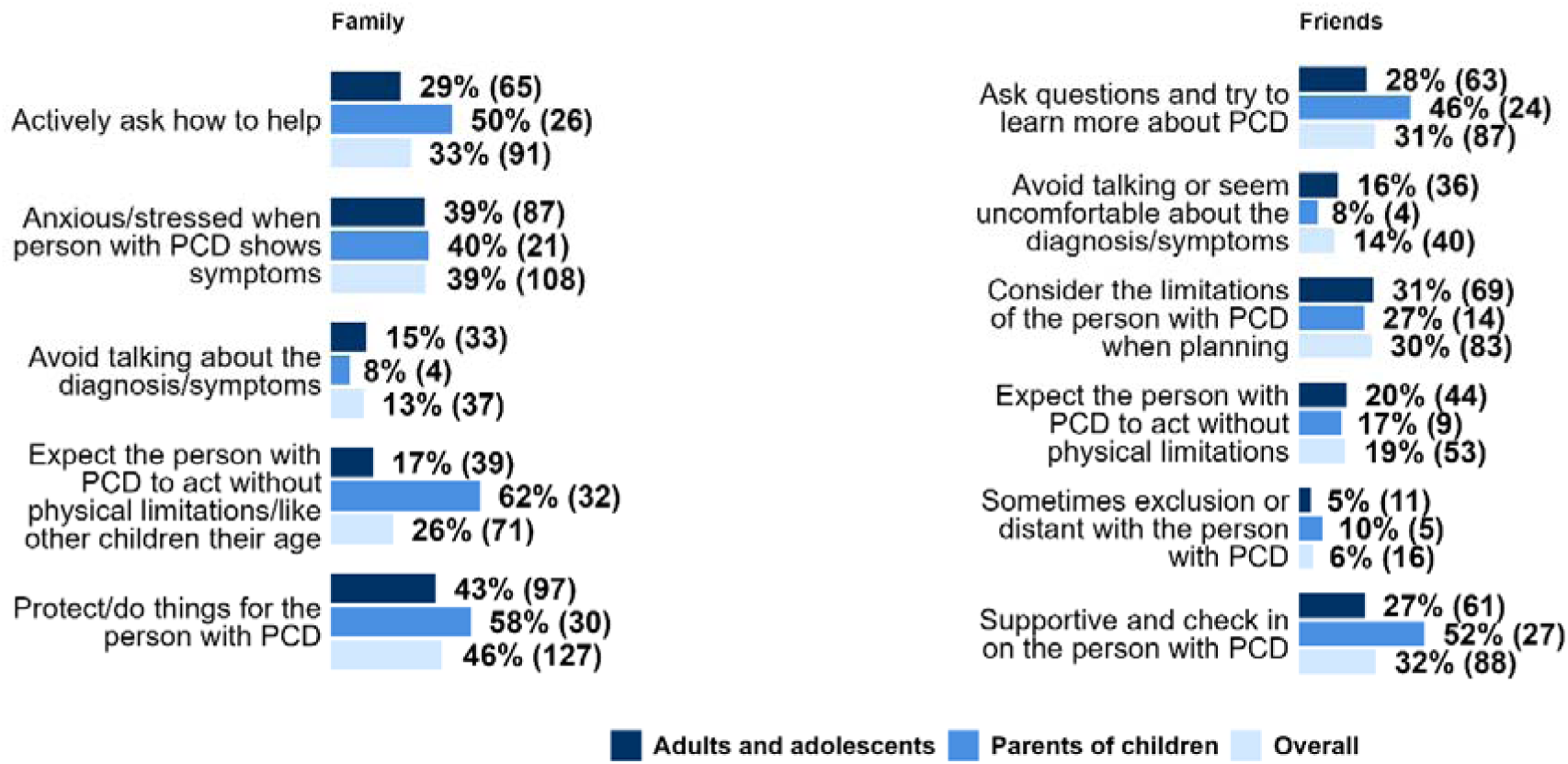
Family and friend’s interpersonal dynamics. Responses from people with primary ciliary dyskinesia registered in the Living with PCD study who completed the social functioning. Additional ways of interaction within the family reported: Support and encourage the person with PCD (n=7), Dismiss the symptoms or expect simple solutions (n=2), Overprotective and controlling (n=1), Lack of interest (n=1). 17% of adults and adolescents (n= 38) and 2% of parents of children with PCD (n=1) reported absence of the listed family dynamics. Additional ways of interaction with friends reported: Lack of understanding or forget about it (n=3), Support and understanding (n=2), Protect the person with PCD (n=1), Downplay PCD importance (n=1), Lack of knowledge about the condition (n=1), Joking about PCD (n=1)”. 24% of adults and adolescents (n= 54) and 15% of parents of children with PCD (n=8) reported absence of the listed friendship dynamics. questionnaire, among parents of children (n=52), adults and adolescents (n=225) and overall (n=277).

### Social involvement and identity and relationship with own body

Participants also reported on broader involvement in social life and their social identity (Table S1). Overall, responses suggested that many participants maintained an active social life and had integrated PCD into their social identity, although some reported that PCD limited participation or shaped how they saw themselves in social contexts. Participants further reported on their relationship with their bodies and physical appearance (Table S2). Many described positive or accepting feelings towards their bodies, while others reported frustration with bodily limitations or appearance-related concerns linked to PCD.

### Management strategies and PCD related experience in different settings

PCD added a substantial burden on daily life, impacting responsibilities, leisure time, and mental effort. Overall, 54% (150) of participants reported that PCD affected their ability to meet responsibilities, 58% (160) reported that it affected their free time, and 53% (149) reported that organising activities required extra planning or mental effort. These impacts were generally reported more often by adults and adolescents with PCD than by parents of children (responsibilities 56% vs. 46%, free time 59% vs. 52% and planning effort 55% vs. 47%, respectively).

To compare the impact of PCD across different life settings, we grouped management strategies and experiences into nine broad categories (see Supplementary Methods for explanation on the grouping). Table 3 summarises the prevalence of these reported experiences at work, education, and leisure/travel settings.

**Table 3.**
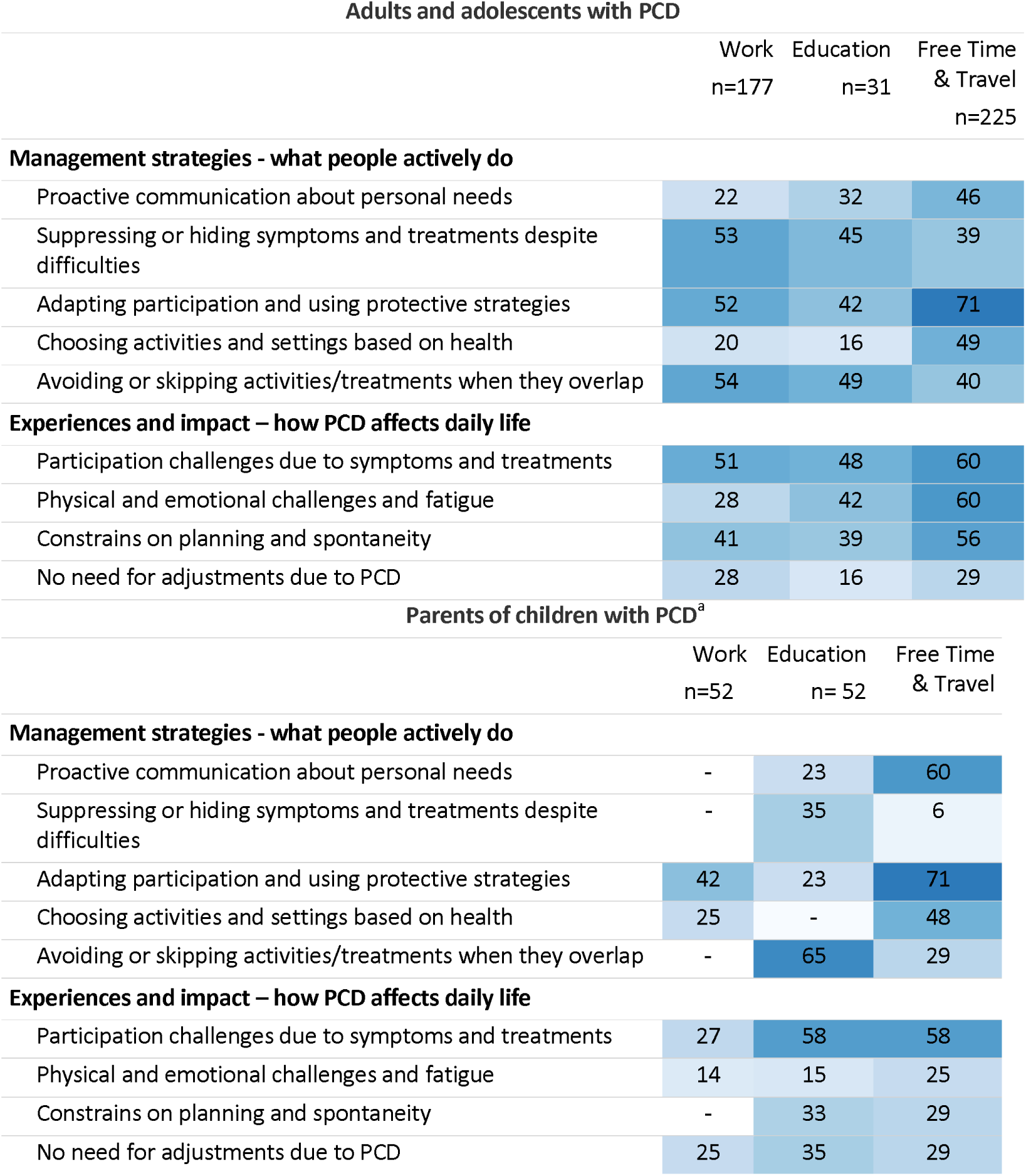
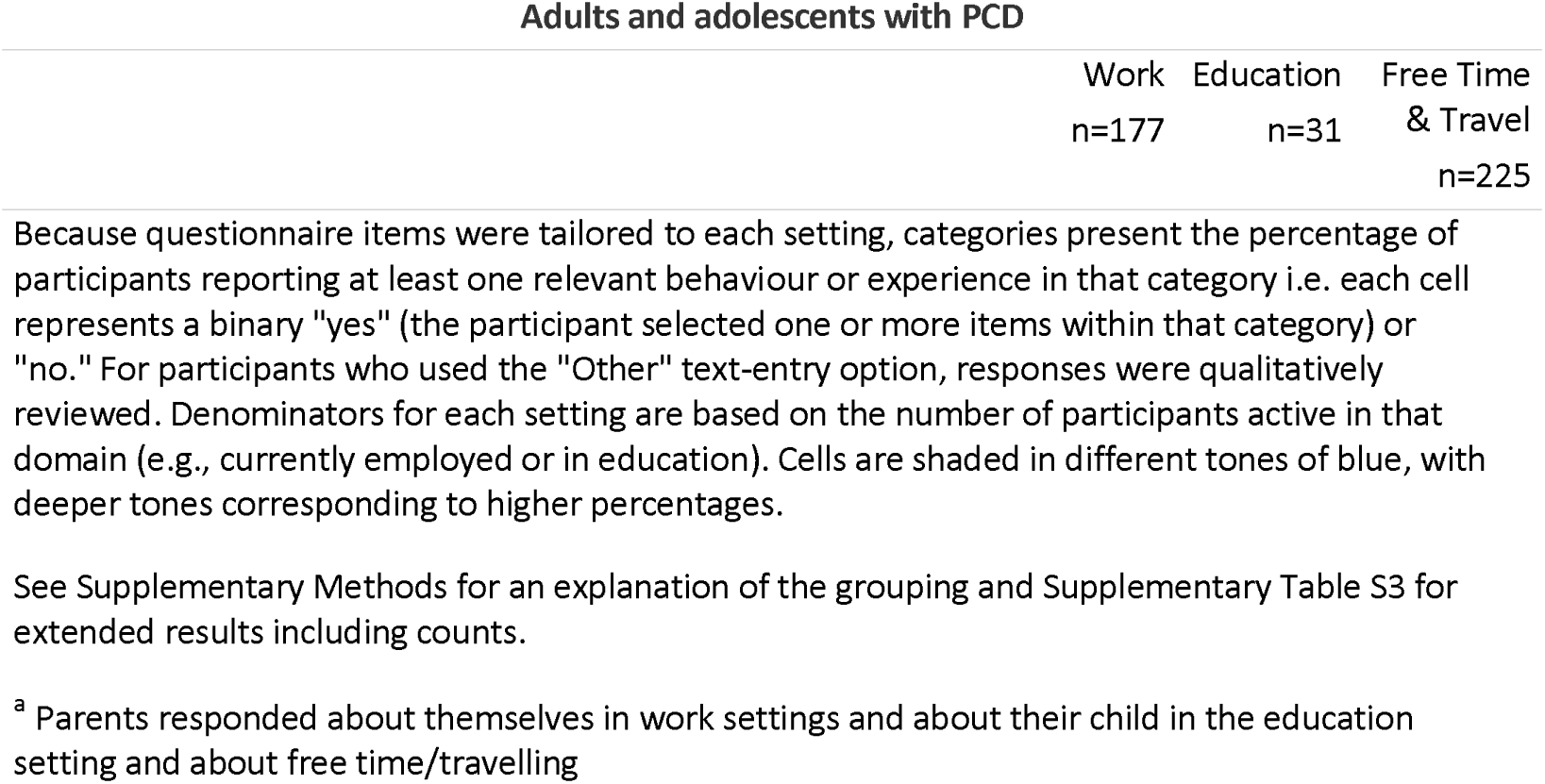
Management strategies and experiences related to PCD across different settings. Responses from people with primary ciliary dyskinesia registered in the Living with PCD study who completed the social functioning questionnaire.

| Adults and adolescents with PCD |  |  |  |
| --- | --- | --- | --- |
|  | Work<br>n=177 | Education<br>n=31 | Free Time<br>& Travel<br>n=225 |
| **Management strategies - what people actively do** |  |  |  |
| Proactive communication about personal needs | 22 | 32 | 46 |
| Suppressing or hiding symptoms and treatments despite difficulties | 53 | 45 | 39 |
| Adapting participation and using protective strategies | 52 | 42 | 71 |
| Choosing activities and settings based on health | 20 | 16 | 49 |
| Avoiding or skipping activities/treatments when they overlap | 54 | 49 | 40 |
| **Experiences and impact – how PCD affects daily life** |  |  |  |
| Participation challenges due to symptoms and treatments | 51 | 48 | 60 |
| Physical and emotional challenges and fatigue | 28 | 42 | 60 |
| Constrains on planning and spontaneity | 41 | 39 | 56 |
| No need for adjustments due to PCD | 28 | 16 | 29 |
| Parents of children with PCD<sup>a</sup> |  |  |  |
|  | Work<br>n=52 | Education<br>n= 52 | Free Time<br>& Travel |
| **Management strategies - what people actively do** |  |  |  |
| Proactive communication about personal needs | - | 23 | 60 |
| Suppressing or hiding symptoms and treatments despite difficulties | - | 35 | 6 |
| Adapting participation and using protective strategies | 42 | 23 | 71 |
| Choosing activities and settings based on health | 25 | - | 48 |
| Avoiding or skipping activities/treatments when they overlap | - | 65 | 29 |
| **Experiences and impact – how PCD affects daily life** |  |  |  |
| Participation challenges due to symptoms and treatments | 27 | 58 | 58 |
| Physical and emotional challenges and fatigue | 14 | 15 | 25 |
| Constrains on planning and spontaneity | - | 33 | 29 |
| No need for adjustments due to PCD | 25 | 35 | 29 |

Adults/adolescents frequently hid symptoms at work or school (95 of 177 working, 53% and 14 of 31 at school, 45%), and adapted activities for leisure and travel (158, 71%). We inquired further about how often adults and adolescents hid and justified their symptoms in public. Nearly half of the 225 adults/adolescents reported always or often justifying or hiding symptoms in public (109, 48%), 28% (63) did so occasionally, and 24% (53) did so rarely or never. Over half of adult/adolescent participants reported that PCD symptoms and treatment demands led to challenges when participating in different settings, most frequently in free time and travel (134, 60%). 65% of parents (34) reported that children skipped treatments or activities due to school overlaps. During leisure time, most parents reported communicating their child’s needs to others (31, 60%) and mentioned the need to adapt participation and using protective strategies (30, 58%). Notably, parents reported lower rates of symptom concealment for their children in social settings (3, 6%) compared to adults/adolescents, who prioritised planning and adapting activities to be able to participate (158, 71%). Additional results on caregiving responsibilities are presented in Table S3, with both groups noting physical and emotional challenges and fatigue (92% of adults/adolescents and 83% of parents).

We additionally asked parents about caregiving for a child with PCD. Very few reported disagreements with their child on treatment management (2, 4%), and slightly more indicated that their child did not contribute to managing their own treatments (5, 10%). One-third reported difficulties in balancing caring for their child with other responsibilities (17, 33%), and in giving equal attention to the child’s siblings (18, 35%). Few parents indicated that the child’s siblings sometimes took on caregiving roles or were impacted by the sibling’s PCD impacted by their sibling’s condition (8, 15%).

### Perceptions, disclosure, and social support needs

Many adults/adolescents (110, 49%) and parents (37, 71%) perceived PCD as an “invisible” condition (Table 4). This invisibility influenced if they disclosed the diagnosis, which varied significantly by social context. Disclosure was common among adults/adolescents’ friends (128, 57%) and among parents’ own friends (43, 83%), as well as within romantic partnerships (153, 68%) of adults/adolescents. In contrast, only 34% (60 out of 177 at work) of adults/adolescents felt comfortable disclosing their diagnosis at work, and similarly low rates were reported for school by 26% of adults/adolescents (8 out of 31 at school) and 29% (15) of parents). Reasons for disclosing or not disclosing having PCD at work or school varied, but the most common reason for not disclosing was that participants considered PCD not relevant in that setting (work: 73, 41%; school: 14, 45%; Table S4). Other reasons included concerns about being treated differently (work: 55, 29%; school: 9, 29%) and fear of stigma (work: 53, 29%; school: 8, 26%). Some participants reported feeling empowered by talking about their diagnosis (work: 34, 18%; school: 5, 16%), needing formal or informal adjustments (work: 23, 13%; school: 5, 16%), or being encouraged by advice from family, friends or professionals (work: 14, 8%; school: 4, 13%).

**Table 4.** Perceived visibility of PCD, disclosure of the diagnosis in different settings, perceived support in different settings and perceived stigma. Responses from reported by people with primary ciliary dyskinesia registered in the Living with PCD study who completed the social functioning questionnaire.

|  | Overall<br>n= 277 | Adults and adolescents<br>n=225 | Parents of children with PCD<br>n=52 |
| --- | --- | --- | --- |
| **Perceived visibility of PCD** |  |  |  |
| Very/ somewhat visible | 97 (35) | 88 (39) | 9 (17) |
| Neutral | 32 (12) | 26 (12) | 6 (12) |
| Very/ somewhat invisible | 148 (53) | 110 (49) | 37 (71) |
| **Diagnosis disclosure to friends** |  |  |  |
| Yes, most of my friends know | 171 (62) | 128 (57) | 43 (83) |
| Yes, only a few close friends know | 83 (30) | 75 (33) | 8 (15) |
| No, I usually don't talk about it | 19 (7) | 18 (8) | 1 (2) |
| No response | 4 (1) | 4 (2) | 0 (0) |
| **Diagnosis disclosure to partner <sup>a</sup>** |  |  |  |
| Very/somewhat comfortable |  | 153 (68) | - |
| Neutral |  | 31 (14) | - |
| Very/somewhat uncomfortable |  | 29 (13) | - |
| No response |  | 12 (5) | - |
| **Diagnosis disclosure at work <sup>ab</sup>** |  |  |  |
|  |  | **n = 177** |  |
| Very/somewhat comfortable |  | 60 (34) | - |
| Neutral |  | 34 (19) | - |
| Very/ Somewhat uncomfortable |  | 42 (23) | - |
| Not informed about PCD |  | 33 (19) |  |
| No response |  | 8 (5) | - |
| **Diagnosis disclosure at school <sup>b</sup>** |  |  |  |
|  | **n=83** | **n=31** | **n=52** |
| Very/somewhat comfortable | 23 (28) | 8 (26) | 15 (29) |
| Neutral | 19 (23) | 7 (23) | 12 (24) |
| Very/ Somewhat uncomfortable | 21 (26) | 5 (16) | 16 (32) |
| Not informed | 10 (12) | 10 (32) | 0 (0) |
| No response | 9 (11) | 1 (3) | 8 (15) |
| **Support by family** |  |  |  |
| Very/somewhat supported | 219 (79) | 170 (76) | 49 (94) |
| Neutral | 30 (11) | 28 (12) | 2 (4) |
| Not really supported/not at all | 23 (8) | 22 (10) | 1 (2) |
| No response | 5 (2) | 5 (2) | 0 |
| **Adjustments/ support at work<sup>ab</sup>** |  | **n=177** |  |
| Very/somewhat supported |  | 51 (29) |  |
| Neutral |  | 27 (15) |  |
| Little support |  | 19 (11) |  |
| No support. I feel they are actively unaccommodating |  | 14 (8) |  |
| Not informed work |  | 33 (19) |  |
| No need extra adjustments |  | 29 (16) |  |
| No response |  | 4 (2) |  |
| **Adjustments/support at school<sup>b</sup>** | **n=83** | **n=31** | **n=52** |
| Very/somewhat supported | 31 (38) | 10 (32) | 21 (41) |
| Neutral | 8 (10) | 4 (13) | 4 (8) |
| Little support | 5 (6) | 1 (3) | 4 (8) |
| No support. They are actively unaccommodating | 3 (4) | 1 (3) | 2 (4) |
| Not informed school | 10 (12) | 10 (33) | 0 (0) |
| No need extra adjustments | 24 (29) | 5 (16) | 19 (38) |
| No response | 1 (1) |  | 1(1) |
| **Support by others with PCD** |  |  |  |
| Very/somewhat | 95 (34) | 73 (33) | 22 (43) |
| Neutral | 16 (6) | 16 (7) | 0 (0) |
| Not really/at all | 7 (3) | 7 (3) | 0 (0) |
| No contact but would like it | 115 (41) | 92 (41) | 23 (43) |
| No contact, wouldn't like it | 41 (15) | 34 (15) | 7 (14) |
| No response | 3(1) | 3(1) | 0 (0) |
| **Better understanding by the PCD community<sup>b</sup>** | **n=121** | **n=99** | **n=22** |
| Agree | 106 (88) | 87 (88) | 19 (86) |
| Neutral | 11 (9) | 9 (9) | 2 (9) |
| Disagree | 3 (3) | 2 (2) | 1 (5) |
| No response | 1 (0) | 1 (1) | 0 |
| **Frequency of stigma experience** |  |  |  |
| Always/ often | 30 (11) | 29 (14) | 1 (2) |
| Occasionally | 87 (31) | 77 (34) | 10 (19) |
| Rarely/ never | 155 (56) | 118 (52) | 37 (71) |
| No response | 5 (2) | 1 (1) | 4 (8) |

Results are presented as count (percentage)
<sup>a</sup> Question not included in age-specific version of the questionnaire completed by parents of children with PCD
<sup>b</sup> A different total n for these questions is specified because they were branched in the questionnaire

Similarly, perceptions of social support reflected disclosure of the diagnosis. While family support was rated highly across participants (219, 79%), reaching 94% (49) among parents of children with PCD, support in formal settings was more limited (Table 4). Only 29% of participants felt supported at work (51 out of 177), and less than half felt supported in school [32% adults/adolescents (10 out of 31); 41% parents (21 out of 52)]. Participants comfortable discussing PCD tended to report more support, while those who were somewhat uncomfortable reported more often little or no support.. In line with the limited perceived support at work, 43% (76 out of 177) of adults/adolescents reported that PCD had affected or threatened their job security (Table S5). Among parents of children with PCD, 13% (7) reported concerns about their own job security. Reported concerns among participants included ability to continue working, frequent absences, negative workplace reactions after diagnosis disclosure, difficulties applying for or keeping a job, and financial consequences such as not being promoted. Among 39 retired participants, 72% (28) reported that they had retired earlier than planned due to PCD.

Regarding peer support, 34% (95) of participants reported feeling supported by others with PCD (Table 4). Although 41% (92) of participants were not currently in contact with other people with PCD, they expressed a desire to form such connections. Among those in contact with other people with PCD, the vast majority (106 out of 121, 88%) agreed that others with PCD understood their experiences better than those without the condition. Experiencing stigma was rare, with 52% (118) of adults/adolescents and 71% (37) of parents stating that they or their children never or rarely experience stigma.

### Areas of unmet support

Participants considered financial support for care (127, 46%), flexibility in work and educational policies (124, 45%), and better-trained health care professionals (123, 44%), as the most important means of assistance they would like to receive (Figure 3).

**Figure 3.**
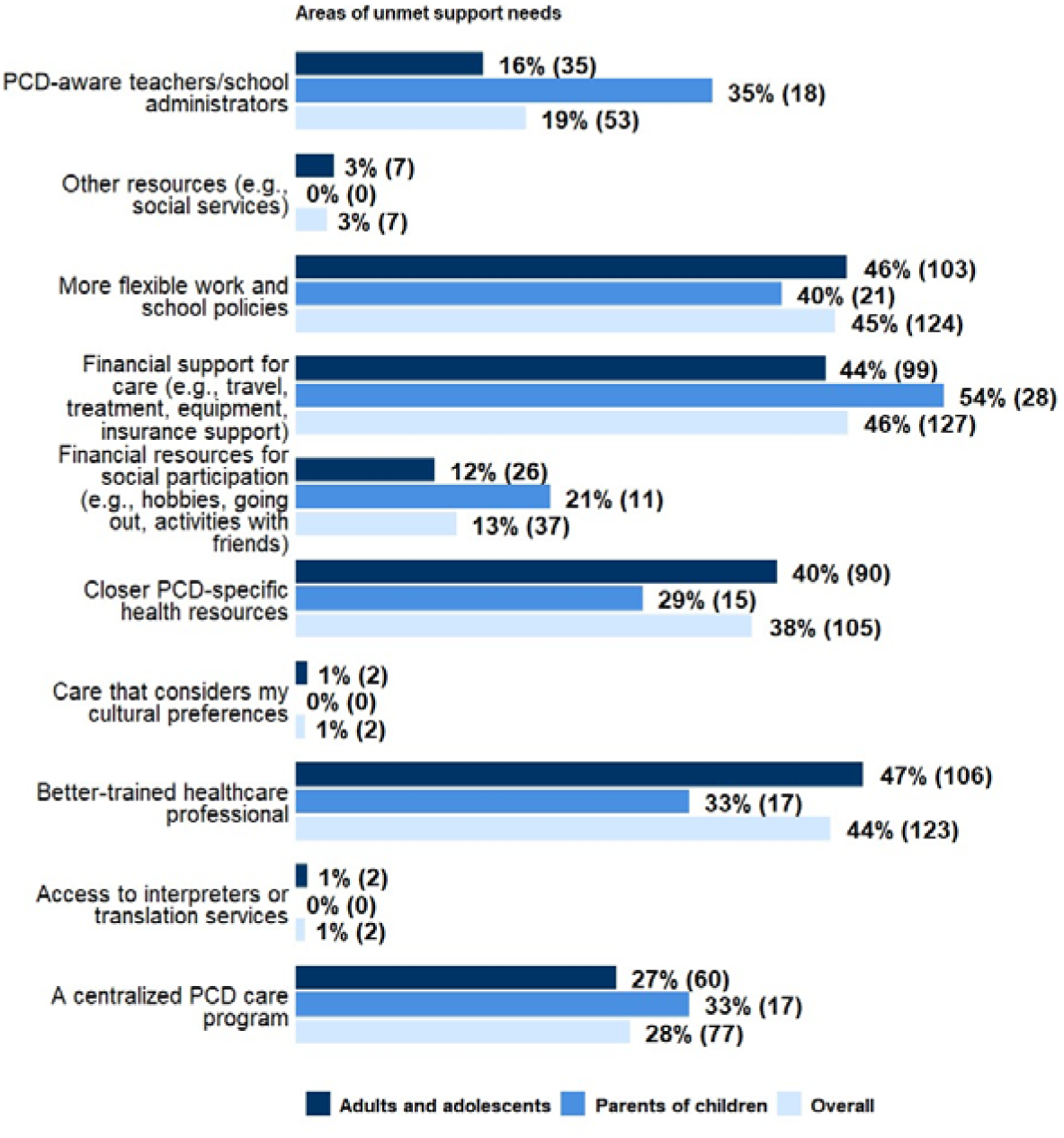
Areas of unmet support reported by people with primary ciliary dyskinesia registered in the Living with PCD study who completed the social functioning questionnaire among parents of children. Additional areas mentioned: Social services to connect to other support (n=1), Mental health Support (n=2), Increased collaboration between health services (n=1), More public awareness (n=1), Support with the household (n=1). (n=52), adults and adolescents (n=225) and overall (n=277)

### Factors associated with communication behaviours in people with PCD

Perceived support at work or education was associated with feeling comfortable discussing PCD among 147 participants. Participants who felt supported in these settings had over two times higher odds of feeling comfortable discussing PCD than those who did not (Odds Ratio-OR: 2.38; 95 % Confidence Interval-CI: 1.18–4.91) (Figure 4). In contrast, we found no signs of association between perceived support at work or education (OR:0.79; 95% CI: 0.37-1.68) or family support (OR:0.60; 95% CI: 0.24–1.51) and hiding or justifying symptoms in public (n=121). We also found no signs of association between age and sex and both outcomes.

**Figure 4.**
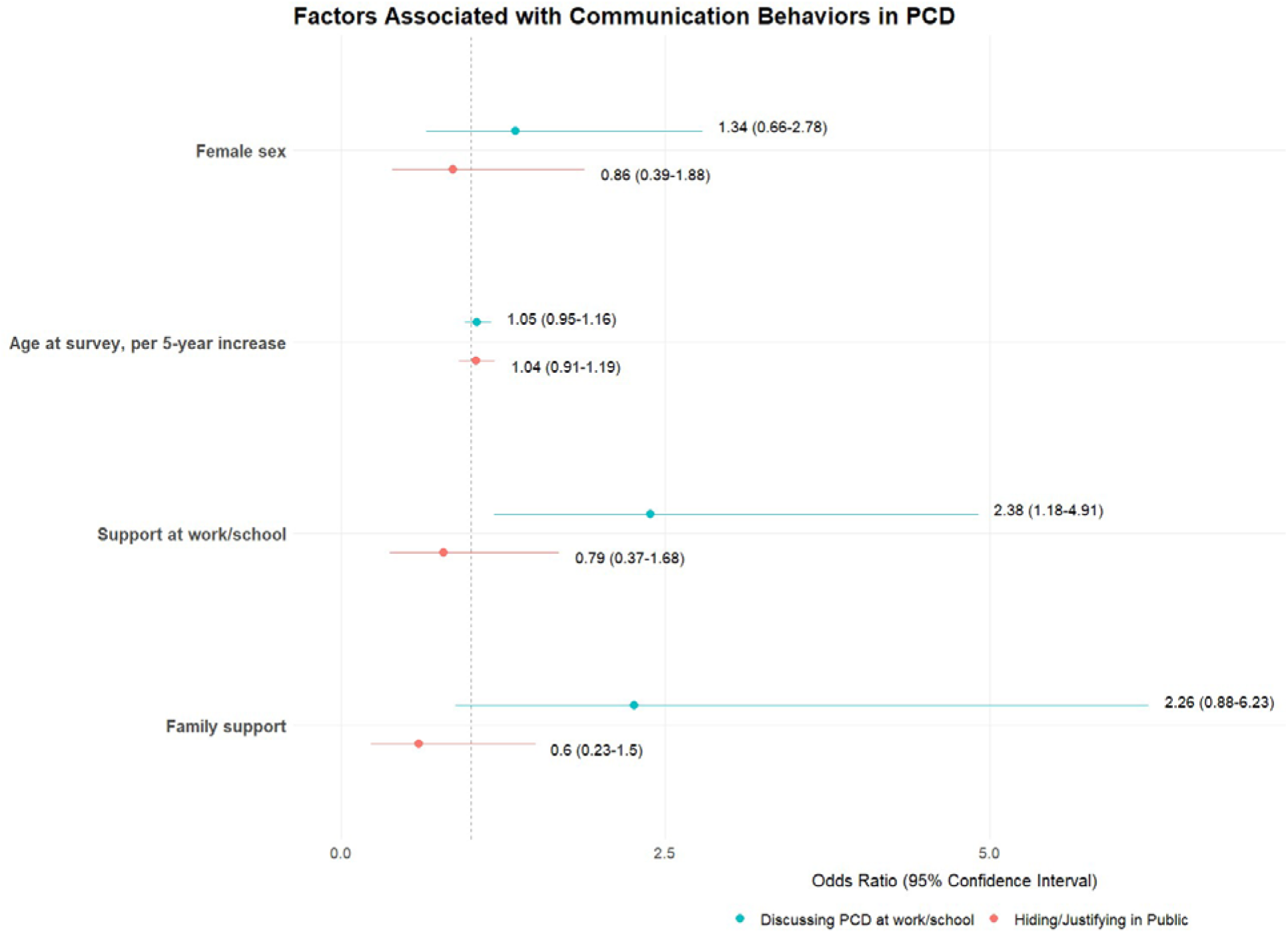
Multivariable logistic regression analysis of factors associated with PCD disclosure and symptom concealment. The forest plot displays potential associations between demographic/support factors and two outcomes: proactive discussion of PCD at work or in an educational setting (blue, n=147) and the tendency to hide or justify symptoms in public (red, n=121). Numerical values represent Odds Ratios (95% Confidence Interval)

## Discussion

This study provides an in-depth exploration of SF in people with PCD. Our findings show that participants often reported active social lives and strong close relationships, yet maintaining participation in different social settings required continuous adaptation, planning, disclosure decisions, and support negotiation. PCD frequently limited participation, increased burden when planning activities, and created challenges in formal settings such as school and work. The difference between private and formal settings was more pronounced when looking at PCD diagnosis disclosure and perceived support, with participants reporting greater openness and support within close relationships than in work or education settings. The areas of unmet support reported by the participants highlighted the need for stronger systemic support such as financial support, flexible work policies, and better trained health care professionals. Support at institutional settings together with support from family were important about disclosing PCD diagnosis at school/work.

In our study, family relationships were often described by people with PCD as strong and a source of support. Our findings are in line with existing studies that showed that family and close relationships provide emotional and practical support in people with chronic respiratory conditions, which can positively influence health outcomes (26). At the same time, this support comes usually with substantial caregiver burden, altered family roles, occupational sacrifices, and broader household disruption, particularly for parents and primary caregivers managing intensive daily treatments as has been shown in CF (27). Interestingly in our study, people with PCD more frequently reported physical and emotional challenges in their own role as caregivers, compared to parents of children with PCD. These findings go beyond usual assumptions that people with PCD are primarily recipients of support, as many occupied caregiving roles themselves, highlighting that SF includes the ability to care for others while managing one’s own symptoms and treatment demands.

Romantic relationships highlighted a different dimension of SF, that of the vulnerability, dependency, and family planning. Participants reported avoiding or ending relationships due to fear of being perceived as burden, fertility concerns, and negative self-perception, consistent with previous findings in PCD (7,28). Research in adults with CF has also highlighted that chronic symptoms, reproductive concerns, and fears of burdening a partner can complicate dating, intimacy, and family-building, while couples often manage these challenges through shared understanding, flexibility and practical support (29). Recent work on reproductive decision-making in CF further shows how rare genetic chronic diseases can shape imagined family life by requiring people to anticipate disease progression, parenting capacity, and the wellbeing of future children (30). These findings highlight romantic relationships as an under-recognised dimension of SF in chronic respiratory diseases. Similar challenges were reported by parents of children with PCD, 42% of whom indicated that the condition had made their own romantic relationship more complicated. Previous qualitative work in parents of children with PCD has highlighted the emotional and practical demands associated with caregiving, but the impact of these demands on parent’s relationships has received little attention (31). However, this finding is consistent with broader research on chronic conditions that report associations between caregiving demands, relationship strain and reduced marital satisfaction (32–34).

Support from others with PCD was identified as an important yet underused resource, which is common for other rare conditions (35). Although many participants were not already in contact with others with PCD, they expressed desire for peer connection, and those with such contact overwhelmingly felt more understood by peers with PCD compared to peers without PCD. Given the rarity of PCD and the frequently experienced lack of awareness and understanding from others, peer support networks, including patient support groups, may offer significant psychosocial benefits by reducing isolation, normalising experiences, and improving coping (35–37).

Most participants reported having integrated PCD into their identity while still maintaining active social lives. Rather than showing social withdrawal behaviours, as has been described in some paediatric populations (38,39), our findings suggest that people incorporate PCD into their sense of self and pursue meaningful social participation. This, however, does not diminish the burden that PCD poses on daily life. Participants described PCD as a constant state of vigilance requiring substantial physical, emotional, and planning effort and highlighted that disease management imposes limitations on work, education, and leisure. Similar findings have been reported in CF where the daily “work” of chronic disease management significantly affects quality of life beyond symptom severity alone (40,41). We hypothesise that this loss of spontaneity may contribute to reduced quality of life in PCD, as SF is shaped not only by physical symptoms (12) but also by the anticipatory work required to participate safely in social life, and that it could be even greater for people with more advanced disease, which is an aspect that requires further research.

Consistent with previous literature describing embarrassment, public self-consciousness, and symptom concealment in PCD (13,42), participants in our study reported actively hiding or concealing symptoms especially in formal settings such as work and school, while few participants reported perceiving stigma from others. The visible–invisible character of PCD created uncertainty: symptoms could be noticeable, but their meaning was often unclear to others. Disclosing the diagnosis was setting-dependant: participants were more comfortable sharing their diagnosis with family and friends than at work or education, and fear of being treated differently was common after disclosing the diagnosis, as reported in other chronic conditions (43,44). In our study, perceived institutional support was associated with feeling comfortable discussing PCD. This suggests that disclosing one’s diagnosis is not an individual choice but rather an informed decision that depends on whether work and school are perceived as safe, informed, and supportive.

Participants frequently mentioned healthcare professionals’ limited knowledge of PCD, and inflexible institutional policies that failed to accommodate treatment demands. Employment insecurity, insurance difficulties, and inadequate social, employment policies further contributed to social disadvantage. These findings emphasise that social challenges in PCD are not only disease-driven but are also reinforced by institutional inequities. Improving SF therefore requires more than psychosocial support for individuals and their families; it requires educating healthcare professionals, visible and well-communicated inclusive practices across healthcare, education and employment, such as flexible school and workplace policies, financial protection, and institutional arrangements, that reduce the burden currently placed on people with PCD and their families. In our study social functioning in PCD is possible not only through individual adaptation but also through supportive relationships. Participants frequently described supportive family relationships, friendships, romantic partnerships and connections with others living with PCD. These relationships appeared to provide emotional support, practical assistance and spaces in which symptoms, treatment demands and uncertainties could be understood and accommodated. Leaning into their community could help people with PCD to increase their social well-being and participate fully in everyday life.

Strengths of our study are the participatory nature of the *Living with PCD* study, which ensures our results are relevant to the PCD patient community, and the online and anonymous study design, which allowed us to involve people with limited involvement in clinical research, from many countries around the world. The design ensured content validity, a complementary approach in a field of research that has largely focused on genetics, diagnosis, and disease physiopathology. Although the qualitative phase involved a relatively small sample, its purpose was to support the development of the questionnaire and it still allowed for in-depth thematic exploration and to reach breadth of information and collect rich and relevant data (45). The main limitation of the study is that it was advertised by patient support groups and was only available in five languages, restricting the population that could be reached, especially in countries with no established support group. It could be possible that people with no contact to a support group face even greater struggles to maintain SF. The relatively small number of parents limited comparisons between participant subgroups and the interpretation of parent-specific findings. Questionnaire responses may also be influenced by selection bias, potentially overrepresenting participants with stronger views on SF or support needs. As our study participants are people with PCD, the social challenges of those undiagnosed or experiencing prolonged diagnosis delays could not be explored, however we hypothesize that people with PCD who remain undiagnosed or receive a diagnosis later in life struggle to maintain SF without even being able to pinpoint the reason and get any support, which highlights the importance of early diagnosis not only for proper medical care but also for all aspects of life. We also did not explore whether social functioning differs based on disease trajectory and severity. Understanding how diagnostic delays and symptom burden may influence social participation could help identify those most at risk of social disadvantage and guide targeted interventions. Although, selection bias cannot be excluded, the presence of both positive and negative accounts of SF suggests that the sample did not exclusively represent participants with unmet support needs. This is further supported by the fact that the frequency of disease-specific characteristics, such as situs abnormalities in our study population is in line with published data. Although our questions focused on current SF, we asked retired participants to report on past work experience, so recall bias may have influenced some responses. However, as our aim was an in-depth exploration of SF, we consider retrospective accounts valuable.

## Conclusion

This is the first study to conduct an in-depth exploration of SF in people with PCD. *Living with PCD* substantially shapes SF through the interaction of disease management, interpersonal relationships, and systemic conditions. People with PCD maintain SF, which requires significant adaptation, negotiation, and advocacy, however the responsibility of doing so currently rests largely on themselves and their close networks. Addressing SF in people with PCD requires healthcare professionals, educational institutions, employers, and policymakers to move beyond individual responsibility towards systemic changes and supportive policies in professional, educational, and health settings. People with PCD are already making substantial efforts to remain socially engaged; it is now essential for systems and institutions to match that effort with meaningful structural support.

## Supporting information

Supplementary methods and results_Social functioning in PCD

## Acknowledgements

We thank all participants and their families, and we thank the PCD support groups, networks, and physicians who advertised the study. We thank our collaborators who helped set up originally the *Living with PCD* study: Cristina Ardura, Yin Ting Lam, Christina Mallet, Helena Koppe, Dominique Rubi, from the University of Bern, but especially, we thank Valérie Schwartz (University of Bern) for her involvement in the current project. We thank the *Living with PCD* advisory group (in alphabetical order): Sara Bellu, Associazione italiana Discinesia Ciliare Primaria Sindrome di Kartagener A.P.S., Italy; Isabelle Cizeau, Association ADCP, France; Fiona Copeland, PCD Support UK, UK; Katie Dexter, PCD Support UK,UK; Lucy Dixon, PCD Support UK, UK; Trini Lopez Fernandez, Asociacion Española de Pacientes con Discinesia Ciliar Primaria/Síndrome de Kartagener, Spain; Susanne Grieder, Selbsthilfegruppe Primare Ciliare Dyskinesie, Switzerland; Catherine Kruljac, PCD Australia, Australia; Michele Manion, PCD Foundation, USA; Hansruedi Silberschmidt, Verein Kartagener Syndrom und Primäre Ciliäre Dyskinesie, Germany: Emilie Wattellier, Association ADCP, France.

## Declaration of generative AI use

To improve the readability and clarity of this manuscript, the authors incorporated grammar and syntax suggestions from Grammarly, DeepL and ChatGPT GPT-5.5 Instant model (web-based free versions). After using this tool, the authors reviewed and edited the content as needed and take full responsibility for the content of this work.

## Author roles

AFR, NK, and MG conceptualised the study. AFR, NK, FP, and MG designed the methodology. All authors contributed to the development of the questionnaire. AFR and NK curated the data. AFR conducted the formal analysis and AFR, NK, and MG drafted the manuscript. KD, MM, and HS contributed patient and public involvement perspectives and supported interpretation of the findings. NK, VG, SZ, FP, CEK and MG contributed to interpretation of the findings. MG supervised the study and acquired funding. All authors critically reviewed and edited the manuscript, approved the final version, and agreed to be accountable for the work.

## Data availability

*Living with PCD* data supporting this manuscript are available upon reasonable request by contacting Myrofora Goutaki.

