## Supplementary methods and results_Social functioning in PCD for "Social functioning in Primary Ciliary Dyskinesia (PCD) – a study of lived experience, relationships and support of patients and caregivers"

The document contains the following:

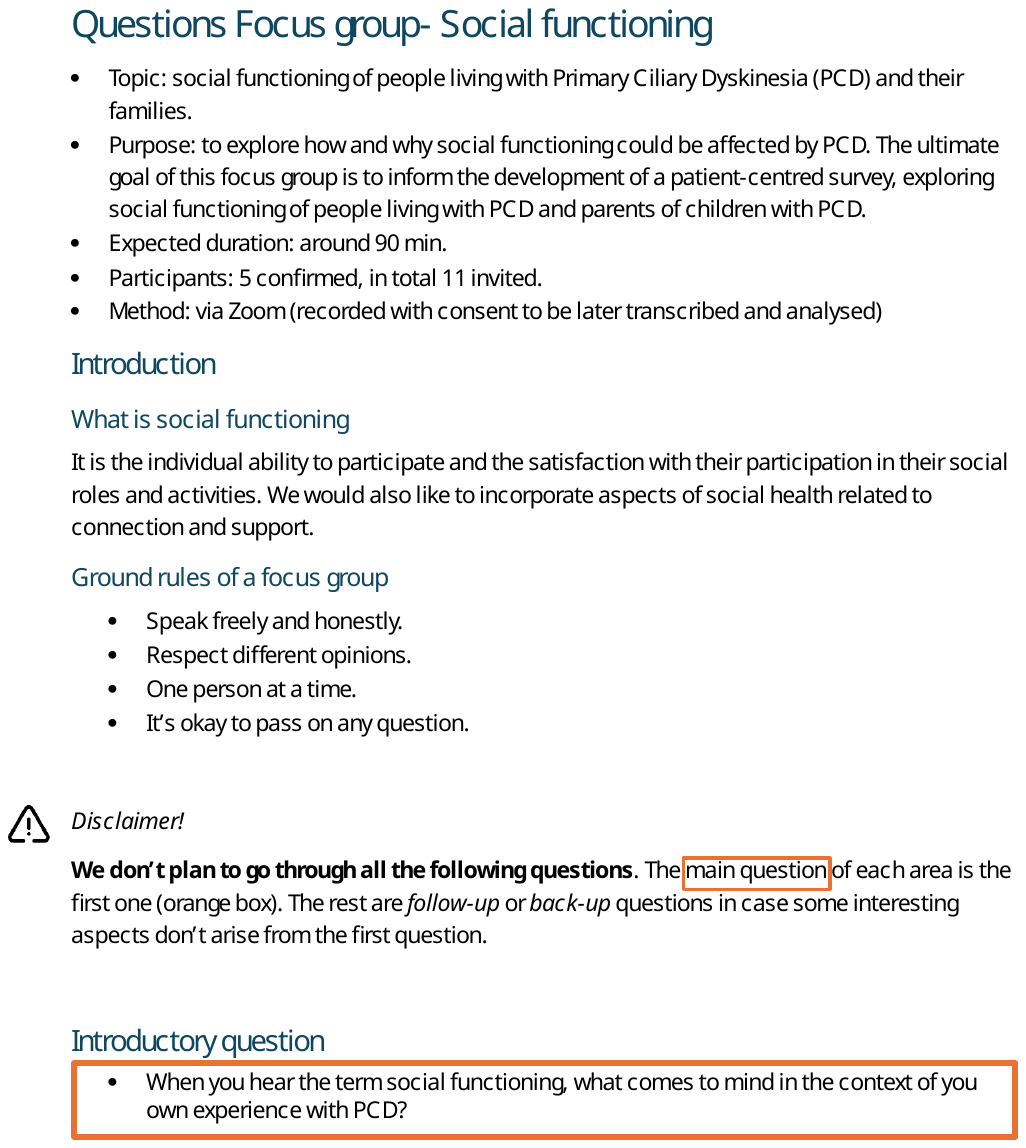

### Focus group guide

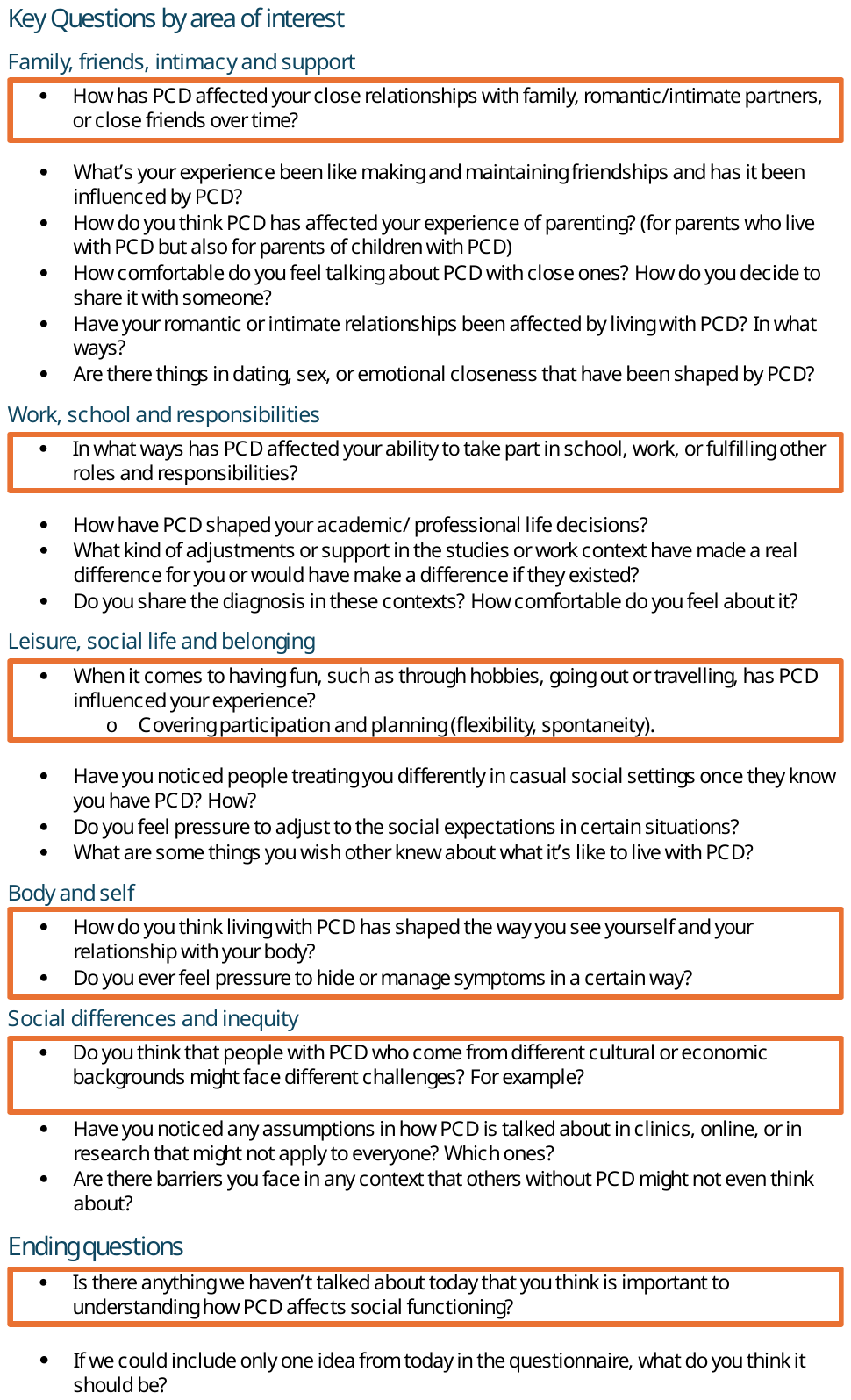

### Social functioning Living with PCD questionnaire: adults’ version

#### Section 1: Basic data

*In the following section, we ask some questions about you to make sure we capture your up-to-date sociodemographic information that might relate to your experiences.*

1. In which country do you live?
2. Did you or your parents move from another country to where you are currently living?
   i. Yes, both my parents and I
   ii. Yes, both of my parents
   iii. Yes, one of my parents
   iv. Yes, myself
   v. No
   vi. I prefer not to answer
3. Is your first language different from the main language(s) spoken in the country you currently live?
   i. Yes
   ii. No
4. Do you have any children?
   i. Yes
   ii. No
5. (if yes) How many of your children are 5 years old or younger?
6. (if yes) How many of your children are between 6 and 17 years old?
7. (if yes) How many of your children are aged 17 or older?

#### Section 2: Relationships

#### Family

8. Has your relationship with your family (e.g., parents, children, siblings) been influenced? *There will be questions about partner(s)/spouse(s) later.*
   i. I think PCD strongly contributed to having strong/close family relationships
   ii. I think PCD somewhat contributed to having strong/close family relationships
   iii. I don’t think PCD has played a big role in the relationship
   iv. I think PCD somewhat contributed to having weak/poor family relationships
   v. I think PCD strongly contributed to having weak/poor family relationships
9. How supported do you feel by your family?
   i. Very supported
   ii. Somewhat supported
   iii. Neutral
   iv. Not really supported
   v. Not supported at all
10. Which of the following apply to your family? (**Tick all that apply**)
    i. They try to protect me or do things for me
    ii. They actively ask how they can help
    iii. They become anxious or stressed when I show symptoms around them
    iv. They expect me to act like I don’t have any (physical) limitations
    v. They avoid talking about my diagnosis/symptoms
    vi. None of the above
    vii. Other (please specify)

#### Friendships

11. Do you tell friends that you have PCD?
    i. Yes, most of my friends know
    ii. Yes, only a few close friends know
    iii. No, I usually don’t talk about it
12. Which of the following apply to your friends who know you have PCD? (**Tick all that apply**)
    i. They consider my limitations (e.g., energy, physical needs) when planning activities
    ii. They ask questions and try to learn more about PCD
    iii. They are supportive and check in on me
    iv. They expect me to act like I don’t have any limitations
    v. They avoid the topic or seem uncomfortable talking about it
    vi. They sometimes exclude me or are distant
    vii. I haven’t really noticed a difference
    viii. Other (please specify)

#### Romantic & Intimate relationships

13. Have your romantic or intimate relationships been influenced? Please, think broadly about relationships with partner(s)/spouse(s), past or present
I. My relationships have been stronger, we have felt closer
II. I don’t think living with PCD has played a big role
III. My relationships have been more complicated
IV. Not applicable, I haven’t had any romantic/intimate relationships
14. Have you ended or avoided a romantic or intimate relationship due to PCD (e.g., due to fertility concerns, symptoms, treatments, concern of burdening them)?
I. Yes
II. No
III. Not applicable
15. (if Q14=yes) If you feel comfortable, please share the reasons in as much detail as you wish.
16. How comfortable are you discussing your PCD diagnosis or health limitations with a partner?
I. Very comfortable
II. Somewhat comfortable
III. Neutral
IV. Somewhat uncomfortable
V. Very uncomfortable
17. How is your experience(s) of emotional closeness and intimacy affected in relationships?
I. Very positively
II. Positively
III. Neutral
IV. Negatively
V. Very negatively

#### Connecting with Others with PCD

18. Are you in contact with others who have PCD?
I. Yes, often
II. Yes, occasionally
III. No, but I would like to
IV. No, I’m not interested
19. (If Q18 = Yes) Does it help?
I. Very much
II. Somewhat
III. Neutral
IV. Not really
V. Not at all
20. (If Q18 = Yes) Do you agree that the PCD community understands you better than those without PCD?
I. Strongly agree
II. Agree
III. Neutral
IV. Disagree
V. Strongly disagree

#### Section 3: Work, school and responsibilities

21. Which of the following apply to you? (**Tick all that apply**)
I. Full-time employment
II. Part-time employment
III. School, university or other education
IV. Retired
V. Caregiving responsibilities (e.g., for children, elderly family)
VI. Other (please specify)
VII. Not currently involved in any of these roles
22. Do you agree that PCD affects your ability to meet your responsibilities?
I. Strongly agree
II. Agree
III. Neutral
IV. Disagree
V. Strongly disagree

#### Professional/work context (if Q21= employment or retired)

23. In what ways do you usually manage symptoms or treatments during work hours? If you are currently retired, please think about your previous work experience. (**Tick all that apply**)
I. I take breaks as needed
II. I inform those around me when I need support
III. I push myself through symptoms even if it’s difficult
IV. I hide symptoms to avoid attention
V. I do not do treatments during work hours
VI. I work from home when possible/needed
VII. Other (please specify)
24. Which of the following do you experience at work? (**Tick all that apply**)
I. I miss days due to symptoms or treatments
II. I miss days due to hospitalisation(s)
III. I need more rest breaks than others
IV. I need to stop working to do my treatments and physiotherapy
V. I struggle to plan or commit (e.g., deadlines, for work events like conferences and retreats) due to health unpredictability
VI. I feel unable to meet others’ expectations
VII. I feel unable to meet my own expectations
VIII. I choose/change work field based on what feels manageable with my symptoms
IX. None of the above
X. Other (please specify)
25. Do you receive any adjustments or support at work that have a positive impact? (e.g., flexible schedule, understanding from your boss/colleagues, opportunities to rest, working from home)
    i. A lot of support
    ii. Some support
    iii. Neutral
    iv. Little support
    v. No support. I feel they are actively unaccommodating
    vi. I have not informed work about PCD
    vii. I do not think I need extra adjustments
26. (Q25 ≠ I have not informed work about PCD) How comfortable are you talking about PCD at work?
I. Very comfortable
II. Somewhat comfortable
III. Neutral
IV. Somewhat uncomfortable
V. Very uncomfortable
27. Which of the following influence your decision to share or not share your PCD diagnosis at work? (**Tick all that apply**)
I. I fear judgment or stigma
II. I am concerned about being treated differently
III. I believe that the diagnosis is not relevant
IV. I need formal or informal adjustments
V. I feel empowered by talking about it openly
VI. I receive advice from family, friends or professionals
VII. Other (please specify)
28. Is your job security influenced?
I. Significantly
II. Somewhat
III. Neutral
IV. No, but I am concerned about it
V. No, and I am not concerned about it
29. (if Q28 ≠ No, and I am not concerned about it) Could you tell us in what way(s)? Share your experience in as much detail as you wish.
30. (if Q21 = Retired) Have your plans or experience with retirement been impacted?
I. I had to retire earlier than planned
II. I had to retire later than planned
III. No impact

Caregiving responsibilities (if Q21= Caregiving)

31. In what ways is your role as a caregiver (e.g., for children, elderly people) affected? (**Tick all that apply**)
    i. I must rely more often on others to help with my caregiving responsibilities
    ii. I adjust activities according to my energy levels/symptoms
    iii. I worry that my condition limits the care I can provide
    iv. My PCD has no effect on my caregiving responsibilities
    v. Other (please specify)
32. Which of the following apply to your caregiving experience? (**Tick all that apply**)
    i. I feel appreciated
    ii. I feel closer to them
    iii. I experience physical exhaustion or fatigue
    iv. I experience difficulty lifting, assisting, or physically supporting those I care for
    v. I need to schedule my treatments around their needs
    vi. I experience stress or emotional strain
    vii. I am concerned about exposing them to infections
    viii. I am concerned about being exposed to infections by them
    ix. I Feel guilty about not being able to do more
    x. None of the above
    xi. Other (please specify)

#### Education (if Q21 = Education)

33. In what ways do you usually manage symptoms or treatments during school hours? (**Tick all that apply**)
I. I take breaks as needed
II. I inform those around me when I need support
III. I push myself through symptoms even if it’s difficult
IV. I hide my symptoms to avoid attention
V. I do not do treatments during school hours
VI. I attend school from home when possible/needed
VII. Other (please specify)
34. Which of the following do you experience at school? (**Tick all that apply**)
I. I miss days due to symptoms or treatments
II. I miss days due to hospitalisation(s)
III. I need more rest breaks than others
IV. I need to leave class to do my treatments or physiotherapy
V. I struggle to plan or commit to events due to health unpredictability (e.g., school trips, school clubs)
VI. I feel unable to meet others’ expectations
VII. I feel unable to meet my own expectations
VIII. I chose/change an education path (e.g., type of high school or university studies, if applicable) based on what feels manageable
IX. None of the above
X. Other (please specify)
35. Do you receive any adjustments or support at school that have a positive impact? (e.g., flexible schedule, understanding from teachers, deadline extension, opportunities to rest, exemptions from activities, at-home education).
I. A lot of support
II. Some support
III. Neutral
IV. Little support
V. No support. They are actively unaccommodating
VI. I have not informed school about PCD
VII. I do not think I need extra adjustments
36. Do you receive health support during school?
I. Yes, regularly a school/university nurse supports me
II. No, I do all treatments by myself
III. Not applicable (e.g., I don’t do treatments during school)
37. (If Q36 ≠I have not informed) How comfortable are you talking about PCD in your school?
I. Very comfortable
II. Somewhat comfortable
III. Neutral
IV. Somewhat uncomfortable
V. Very uncomfortable
38. Which of the following influence your decision to share or not share your PCD diagnosis? (**Tick all that apply**)
I. I fear of judgment or stigma
II. I am concerned about being treated differently
III. I believe that the diagnosis is not relevant
IV. I need formal or informal adjustments
V. I feel empowered by talking about it openly
VI. I receive advice from family, friends or professionals
VII. Other (please specify)

#### Section 4: Free time

39. In what ways do you usually manage symptoms or treatments during your free time? (**Tick all that apply**)
I. I openly explain to people what is going on and manage them
II. I warn others in advance
III. I try to hide my symptoms or delay my treatments
IV. I avoid social events that I consider not worth risking my health (e.g., crowded, indoor spaces)
V. I adapt the activity (e.g., leave early, take rest breaks, change to an open-air setting)
VI. I take extra protective measures to avoid infections or any worsening of symptoms (e.g., wearing masks, hand sanitiser)
VII. None of the above
VIII. Other (please specify)
40. How affected is the amount of time you can spend on free time activities (like hobbies or socialising)?
    i. Significantly
    ii. Somewhat
    iii. Neutral
    iv. Not much
    v. Not at all
41. Which of the following do you experience when planning free time or social activities? (**Tick all that apply**)
I. I need to plan around treatments or symptoms
II. I worry about energy levels
III. I need to cancel plans last-minute
IV. I am not able to plan spontaneously
V. I experience difficulties keeping up physically
VI. I have to explain my health to others
VII. None of the above
VIII. Other (please specify)
42. Do you invest extra planning or mental effort to organise your free time?
I. Significantly
II. Somewhat
III. Neutral
IV. Not much
V. Not at all
43. Which of the following **best** describes your current social life?
I. I socialise regularly and feel included
II. I socialise but feel like I have to make extra effort to keep up with my peers
III. I often feel left out or excluded
IV. I don’t have an active social life. I am happy about it
V. I don’t have an active social life. I am unhappy about it
44. In what ways is your travelling affected? (**Tick all that apply**)
I. I avoid travelling long distances
II. I pack a lot more than others (including medications, treatment equipment)
III. I plan my schedule carefully to accommodate treatment and rest time
IV. I travel less often than I would like to
V. I select my travel destinations in relation to my health (e.g., avoid remote locations, avoid locations where I think healthcare is suboptimal or expensive)
VI. I get a medical check-up before travelling
VII. I do an extensive search of the medical options in my destination before the trip
VIII. I select particular travel methods (e.g., avoiding the underground).
IX. My travelling has not been affected
X. Other (please specify)

#### Section 5: How others see you and how you see yourself

#### Others’ perceptions

45. How visible do you think your condition is to others?
    i. Very visible
    ii. Somewhat visible
    iii. Neutral
    iv. Somewhat invisible
    v. Mostly invisible
46. Which of the following helps you feel included or understood socially? (**Tick all that apply**)
i. My family and partner(s) who know about my PCD diagnosis
ii. Friends or peers who know about my PCD diagnosis
iii. Connecting with others with PCD or other chronic conditions
iv. Online or local support groups
v. Adapting how I communicate about my health
vi. Choosing close friends who are understanding
vii. None of the above
viii. Other (please specify)
47. How often do you need to justify/try to hide your symptoms in public (e.g., “just a cold”, “bad asthma”)?
    i. Always
    ii. Often
    iii. Occasionally
    iv. Rarely
    v. Never
48. How often do you experience stigma or negativity from others?
    i. Always
    ii. Often
    iii. Occasionally
    iv. Rarely
    v. Never

#### Self-perception

49. How does living with PCD affect the way you see yourself in social situations and relate to your PCD diagnosis? (**Please choose the statement that BEST describes your experience**)
I. I feel it is a big part of who I am, and it strongly affects how I see myself in social settings
II. I accept it is part of me, and it influences how I interact with others sometimes
III. I know I have it, but I don’t think about it much, and it doesn’t affect how I see myself socially
IV. I don’t feel it plays a role in how I see myself socially
I. I’m still figuring out what the PCD diagnosis means for me and how it fits into my life socially
50. Which of the following describe the relationship with your body? (**Tick all that apply**)
I. I am proud of what my body handles and achieves
II. I feel mostly positive and accepting
III. I have mixed feelings about my body
IV. I feel disconnected from my body
V. I have appearance-related concerns
VI. I am frustrated with limitations
VII. I am ashamed of my symptoms
VIII. I actively avoid mirrors or body-related thoughts
IX. None of the above
X. Other (please specify)

#### Section 6: Everyday challenges and support

51. Which of the following supports would you consider most helpful? (**Tick up to 3**)
    i. Financial support for care (e.g., travel, treatment, equipment, insurance support)
    ii. Financial resources for social participation (e.g., hobbies, going out, activities with friends)
I. More flexible work and school policies
II. Better-trained healthcare professional
III. PCD-aware teachers/school administrators
IV. Care that considers my cultural preferences
V. Closer PCD-specific health resources
VI. Access to interpreters or translation services
VII. A centralised PCD care program
VIII. Other resources (e.g., social services) (please specify)

### Grouping of reported management strategies and PCD related experience in different settings (Tables 3 and S3 construction description)

In our social functioning questionnaire, we developed questions for each life setting (i.e., work, education, caregiving, free time, and travel) to reflect the unique aspects we identified during the focus group discussion and one-to-one interviews. As a result, the same items were not repeated across all settings; questions were adapted to the context of each setting. To facilitate comparison and present findings in the most relevant way, we performed a thematic mapping exercise, grouping setting-specific questions into nine overarching categories based on the core constructs of social functioning in PCD. Taking the adult version as reference (page 4-10 in this document), these questions’ numbers were 23 and 24 related to work, 31 and 32 related to caregiving responsibilities, 33 and 34 related to education, and 40, 42 and 45 related to free time and travel.

**Overarching categories and short definitions**. The following Table details the 9 overarching categories used to present the results in Table 3 and Table S3 and the statements grouped within each of them. Within brackets we include the section of the questionnaire to which those statements belong: work W) or education (school) setting (S), caregiving (C), free time or travel (FT-T).

| **Category** | **Category name** | **What is included** |
| --- | --- | --- |
| **1** | “**Proactive communication about personal needs related to PCD”** | This category includes statements in which participants actively communicate their health needs to others in order to manage symptoms or treatments. This includes informing others when support is needed, explaining their condition, or warning others in advance.  Specific statements   - I inform those around me when I need support (W,S) - I openly explain to people what is going on and manage my symptoms (FT-T) - I warn others in advance (FT-T) - I have to explain my health to others (FT-T) |
| **2** | **“Suppressing or hiding symptoms and treatments despite difficulties”** | This category includes all statements referring to efforts to conceal symptoms or continue activities despite physical discomfort. Participants may push themselves through symptoms or delay treatments, often to avoid attention or disruption.  Specific statements   - I push myself through symptoms even if it’s difficult (W,S) - I try to hide my symptoms or delay my treatments (W,S, FT-T) - Adapting participation and using protective strategies I take breaks as needed (W,S) - I work from home when possible/needed (W,S) - I adapt the activity (e.g., leave early, take rest breaks, change to an open-air setting) (FT-T) - I take extra protective measures to avoid infections or any worsening of symptoms (e.g., wearing masks, hand sanitiser) (FT-T) - My child is exclusively home-schooled (S) - I have reduced my total working hours (W) - I have increased my total working hours (W) - I adjust activities according to my energy levels/symptoms/ I often need to adjust the family routines to manage treatments or appointments (C) - I pack a lot more than others (including medications, treatment equipment) (FT-T) |
| **3** | **Adapting participation and using protective strategies** | This category includes adjustments made to enable continued participation while managing health. Strategies involve modifying activities, schedules, or environments (e.g., taking breaks, working/studying from home, adjusting routines, or bringing additional equipment).  Specific statements   - I take breaks as needed (W,S) - I work from home when possible/needed (W) - I attend school from home when possible/needed (S) - I adapt the activity (e.g., leave early, take rest breaks, change to an open-air setting (FT-T) - My child is exclusively home-schooled (S) - I have reduced my total working hours (W) - I have increased my total working hours (W) - I adjust activities according to my energy levels/symptoms/ I often need to adjust the family routines to manage treatments or appointments (C) |
| **4** | **Choosing activities and settings based on health** | This category captures longer-term or structural decisions shaped by health considerations. Participants may select or modify work, education, travel destinations, or modes of transport based on what is manageable or safe.  Specific statements   - I choose/change an education path based on what feels manageable (W,S) - I select my travel destinations in relation to my health (e.g., avoid remote locations, avoid locations where I think healthcare is suboptimal or expensive) - I avoid travelling long distances - I select particular travel methods (e.g. avoiding the underground) - I have changed jobs to have more flexibility (W) - I avoid social events that I consider not worth risking my health (e.g. crowded, indoor spaces) (FT-T) - I do not do treatments during school/work hours (W,S) |
| **5** | **Avoiding or skipping activities/treatment when they overlap** | This category reflects situations where participants prioritise one demand over another when conflicts arise. This may include skipping treatments during work or school, or avoiding social activities perceived as too risky.  Specific statements   - I do not do treatments during work/school hours - I avoid social events that I consider not worth risking my health (e.g., crowded, indoor spaces) |
| **6** | **Participation challenges due to symptoms and treatments** | This category describes direct disruptions to daily roles and activities caused by symptoms or treatment demands. These include missing work or school, needing to leave activities, reducing travel, or relying on others for support.  Specific statements   - I need to plan around treatments or symptoms (FT-T) - I miss days due to symptoms or treatments (W, S) - I miss days due to hospitalisation(s) (W,S) - I need to leave class to do my treatments or physiotherapy (W,S) - I travel less often than I would like to (FT-T) - I must rely often on others to help with my caregiving responsibilities/ I rely more on support from family, friends to her with my caregiving responsibilities (C) - I experience difficulty lifting, assisting, or physically supporting those I care for (C) - I have frequently taken time off (W) - I have stopped working because of caregiving demands (W) |
| **7** | **Physical and emotional challenges and fatigue** | This category includes the physical burden of PCD alongside its emotional (negative) impact. Participants report fatigue, difficulty keeping up physically, stress, and concerns about their ability to fulfil roles or care for others.  Specific statements   - I worry about energy levels (FT-T) - I experience difficulties keeping up physically (FT) - I need more rest breaks than others (W, S) - I experience physical exhaustion or fatigue (C) - I worry my child misses out on activities because of their condition (C) - I worry that my condition limits the care I can provide (C) - I am concerned about being exposed to infections by them (C) - I am concerned about exposing them to infections (C) - I experience stress or emotional strain (C) - I feel guilty about not being able to do more (C) - I have worried about job security (W) |
| **8** | **Constraints on planning and spontaneity** | This category reflects the impact of unpredictability and the need for constant planning. Participants report difficulty committing to plans, cancelling activities, or being unable to act spontaneously due to fluctuating health. It also includes anticipatory behaviours such as extensive planning to manage potential risks.  Specific statements   - I need to cancel plans last minute (FT-T) - I am not able to plan spontaneously (FT-T) - I struggle to plan or commit to events due to health unpredictability (e.g., school trips, school clubs) (W,S) - I feel unable to meet others expectations (W,S) - I feel unable to meet my own expectations (W,S) - I do an extensive search of the medical options in my destination before the trip (FT-T) - I get a medical check-up before travelling (FT-T) - I do an extensive search of the medical options in my destination before the trip (FT-T) |
| **9** | **No need for adjustments due to PCD** | This category includes participants reporting no perceived impact of PCD on specific areas of life. It reflects either absence of limitations or effective management strategies that allow individuals to function without additional adjustments.  Specific statements   - None of the above (W,S,FT-T) - My PCD has no effect on my caregiving responsibilities/ It doesn't significantly affect how I parent or manage daily life (C) - My travelling has not been affected (FT-T) - Work has not been affected (W) |

SUPPLEMENTARY RESULTS

Social functioning in Primary Ciliary Dyskinesia (PCD) – a study of lived experience, relationships and support of patients and caregivers Andrea Fernandez-Rodriguez, Nena Karavasiloglou, Vasiliki Gkatzou, Katie Dexter, Michele Manion, Hansruedi Silberschmidt, Sofia Zambrano, Francesco Pagnini, Claudia E. Kuehni, Myrofora Goutaki

The document contains the following:

Phase 1: Focus group discussion and one-to-one interviews……………………………………………… 16

**Phase 1: Focus group discussion and one-to-one interviews**

**Theme 1: Having a PCD diagnosis shapes social life**

This theme captures both the personal and social implications of living with a PCD diagnosis, and how it can become a key factor influencing relationships, expectations and identity. Participants described efforts to resist the “role of a sick person,” especially during adolescence, often overcompensating to avoid missing out on social life. Disclosure of the diagnosis was described as an ongoing process rather than a single act: participants weighed when, how, and to whom to explain PCD, while anticipating whether others would recognise their symptoms as legitimate, minimise them, or be open to provide support. In this context, receiving a definite diagnosis was often empowering as it provided the legitimacy needed to inform others.

“Before my diagnosis I always couldn’t really say what’s (wrong) about me… afterwards, it changed totally… I could inform” (P2, patient).

PCD also influences both social expectations and one’s own expectations of themselves. For people with PCD social participation requires constant weighing of wish to participate versus health consequences.

**Theme 2: Everyday management and loss of spontaneity**

Living with PCD requires constant vigilance, which becomes a “default mode.” Participants described being routinely alert to environmental risks and infectious threats. Health-related preparations, such as airway clearance before social events, leave little room for spontaneity in everyday life.

“You can go out... but before you go, you have to do your physiotherapy” (P4, mother).

These affect reciprocity in relationships and influence major life decisions, such as career paths and relocation, which are often dictated by people’s proximity to specialised care.

“I'm hesitating to go back to where I used to work... because I don't know if I will find a good physiotherapist” (P3, patient).

**Theme 3: Support and relationship dynamics**

PCD impacts the entire family unit often resulting in shifts in dynamics, such as parents having to stop working or attention imbalances between siblings. A recurring concern was the perceived lack of validation from relatives and institutions due to the condition’s invisibility. This forced some participants to constantly explain and justify their limitations, reducing their sense of legitimacy.

“Sometimes she's not able to participate... because (otherwise) she has to miss her airway clearance before lunch” (P6, mother).

While some friendships faded over time others proved resilient and support from others with PCD offered meaningful opportunities to create strong bonds.

**Theme 4: System structures and inequities**

Interactions with the healthcare system were frequently frustrating, with participants feeling unheard or mistreated by professionals who lacked knowledge about PCD. Participants considered the use of terms like “mild” to characterise PCD as minimising the real-life impact of the disease. Structural barriers extended to education and employment, where a lack of sick leave or difficulty obtaining insurance created instability.

“When my eldest child tried to get insurance, they wouldn't give him insurance because he's got PCD” (P4, mother).

On the contrary, institutional flexibility, such as adjustable work schedules, was highlighted as a rare but vital support for social participation.

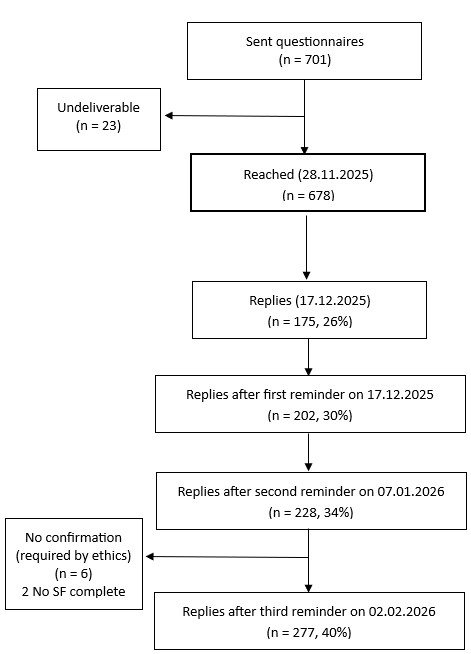

Figure S 1. Flowchart of those registered in the Living with PCD study who completed the social functioning questionnaire

Textbox S 1. Main topics and illustrative comments regarding ending or avoiding romantic or intimate relationships.^1^ Responses from people with primary ciliary dyskinesia registered in the Living with PCD study who completed the social functioning questionnaire

**Concerns about being a burden to the partner**

“I feel it's a lot easier to deal with the limitations and time constraints of my condition being single and it reduces the potential negative impact on someone else.” (Female, 41-60y)

“Until my early 20s, I avoided romantic relationships altogether due to not wanting to burden others with my PCD.” (Male, 21-40y)

**Fertility-related concerns**

“Ended marriage due to not been able to have children.” (Male, >60y)

“My ex-husband and I experienced significant marital distress when we were unable to conceive. He refused to get tested which made me feel so alone in the fertility challenges.” (Female, 21-40y)

**Lack of support from the partner**

“I did not feel they were supportive in any way or wanted to understand the condition.” (Female, >60y)

“I had a husband, but it was hard to cope with the illness during our marriage. He knew what I had, but he didn’t understand the things I couldn’t do; I felt embarrassed to tell him I couldn’t.” Female, >60y)

**Negative self-perception related to symptoms**

“Coughing and a runny nose all day every day isn't really attractive”. (Male, 21-40y)

“I was not diagnosed with PCD until I was in my early 50's. So I did not know what I had I just always felt I had to settle for less because I was snotty and always coughing”. (Female, >60y)

^1^Quotes were selected to illustrate recurrent topics across adults/adolescents respondents who reported having ended or avoided a relationship due to PCD. No formal qualitative analysis was conducted.

Table S 1.Social life involvement and social identity in relation with PCD. Responses from people with primary ciliary dyskinesia registered in the Living with PCD study who completed the social functioning questionnaire, overall and by age group

|  | Overall  n= 277 | Adults and adolescents  n=225 | Parents of children with PCD  n=52 |
| --- | --- | --- | --- |
| Social life involvement |  |  |  |
| Socialise regularly and feel included | 131 (47) | 97 (43) | 34 (65) |
| Socialise but have to make extra effort to keep up with peers | 73 (26) | 62 (28) | 11 (21) |
| Often feel left out or excluded | 11 (4) | 9 (4) | 2 (4) |
| No active social life and happy about it | 38 (14) | 34 (15) | 4 (8) |
| No active social life and unhappy about it | 24 (9) | 23 (10) | 1 (2) |
| Relationship with PCD ^a^ |  |  |  |
| It is a big part of who I am, it strongly affects how I see myself socially | - | 56 (25) | - |
| It is part of me and influences how I interact with others sometimes | - | 88 (39) | - |
| I have it, but I don’t think about it much | - | 62 (28) | - |
| Doesn’t play a role in how I see myself socially | - | 16 (7) | - |
| Still figuring out what the diagnosis means for me and how it fits social life | - | 3 (1) | - |
| Results are presented as counts (n) and column percentages (%)  ^a^Asked only to adult and adolescent participants | | | |

Table S 2. Self-reported relationship with own body in adults and adolescents with primary ciliary dyskinesia registered in the Living with PCD study who responded the social functioning questionnaire

| **Characteristic** | **Adults and adolescents**  n = 225 |
| --- | --- |
| I am proud of what my body handles and achieves | 82 (36) |
| I feel mostly positive and accepting | 94 (42) |
| I have mixed feelings about my body | 87 (39) |
| I feel disconnected from my body | 15 (7) |
| I have appearance-related concerns | 40 (18) |
| I am frustrated with limitations | 92 (41) |
| I am ashamed of my symptoms | 54 (24) |
| I actively avoid mirrors or body-related thoughts | 14 (6) |
| None of the above | 2 (1) |
| Results are presented as counts (n) and column percentages (%) |  |

**Management and experiences across settings**. Participants reported on management strategies and experiences across work, education, free time and travel, and caregiving responsibilities (Table 3 and S3). Caregiving responsibilities are presented only in the supplementary Table S3 because the questionnaire included caregiving-specific items, which did not align with all categories used. Physical and emotional challenges and fatigue related to caregiving were common in both groups but were reported more often by adult/adolescent participants with PCD compared to parents reporting about their own caregiving experiences with a child with PCD (92% vs. 83%). Among adults and adolescents with PCD, 46% also reported that symptoms and treatments made caregiving more difficult.

Table S 3. Extended version of table 3 about Management strategies and experiences related to PCD, across different settings including caregiving responsibilities. Responses from people with primary ciliary dyskinesia registered in the Living with PCD study who completed the social functioning questionnaire

| **Adults and adolescents with PCD** | | | | | | | | | | | | |
| --- | --- | --- | --- | --- | --- | --- | --- | --- | --- | --- | --- | --- |
|  | | Work  n=177 | | Education  n=31 | | | | Caregiving  n=26 | | Free Time & Travel  n=225 | | |
| **Management strategies - what people actively do** | | | | | | | | | | | | |
| Proactive communication about personal needs | | 22  (38) | | | 32  (10) | | | | - | | | 46  (132) |
| Suppressing or hiding symptoms and treatments despite difficulties | | 53  (95) | | | 45  (14) | | | | - | | | 39  (88) |
| Adapting participation and using protective strategies | | 52  (92) | | | 42  (12) | | | | 62  (16) | | | 71  (158) |
| Choosing activities and settings based on health | | 20  (35) | | | 16  (5) | | | | - | | | 49  (109) |
| Avoiding or skipping activities/treatments when they overlap | | 54  (95) | | | 49  (15) | | | | - | | | 40  (88) |
| **Experiences and impact – how PCD affect daily life** | | | | | | | | | | | | |
| Participation challenges due to symptoms and treatments | | 51  (90) | | | | 48  (15) | | | 46  (12) | | | 60  (134) |
| Physical and emotional challenges and fatigue | | 28  (50) | | | | 42  (13) | | | 92  (24) | | | 60  (135) |
| Constrains on planning and spontaneity | | 41  (72) | | | | 39  (12) | | | - | | | 56 |
| No need for adjustments due to PCD | | 28  (49) | | | | 16  (5) | | | 8  (2) | | | 29  (65) |
| **Parents of children with PCD**^a^ | | | | | | | | | | |  | |
|  | Work  n=52 | | Education  n=52 | | | | Caregiving  n=52 | | | | Free Time & Travel  n=52 | |
| **Management strategies - what people actively do** | | | | | | | | | | | | |
| Proactive communication about personal needs | - | | 23  (12) | | | | - | | | | 60  (31) | |
| Suppressing or hiding symptoms and treatments despite difficulties | - | | 35  (18) | | | | - | | | | 6  (3) | |
| Adapting participation and using protective strategies | 42  (20) | | 23  (11) | | | | 67  (35) | | | | 71  (37) | |
| Choosing activities and settings based on health | 25  (12) | | - | | | | - | | | | 48  (25) | |
| Avoiding or skipping activities/treatments when they overlap | - | | 65  (34) | | | | - | | | | 29  (15) | |
| **Experiences and impact – how PCD affect daily life** | | | | | | | | | | | | |
| Participation challenges due to symptoms and treatments | 27  (12) | | 58  (29) | | | | 23  (12) | | | | 58  (30) | |
| Physical and emotional challenges and fatigue | 14  (8) | | 15  (7) | | | | 83  (42) | | | | 25  (13) | |
| Constrains on planning and spontaneity | - | | 33  (16) | | | | - | | | | 29  (25) | |
| No need for adjustments due to PCD | 25  (13) | | 35 | | | | 19  (10) | | | | 29 | |

Participants reporting at least one relevant behaviour or experience in that category i.e. each cell represents a binary "yes" (the participant selected one or more items within that category) or "no." For participants who used the "Other" text-entry option, responses were qualitatively reviewed. Denominators for each setting are based on the number of participants active in that domain (e.g., currently employed or in education). Cells are shaded in different tones of blue, with deeper tones corresponding to higher percentages.^a^ Parents responded about themselves in work settings and caregiving responsibilities, but about their child in the education setting and free time/travelling.

Table S 4. Reasons that facilitate and limit the disclosure PCD at work/school. Responses from adults and adolescents with primary ciliary dyskinesia registered in the Living with PCD study who completed the social functioning questionnaire

| **Facilitators and barriers for disclosure of PCD** | **At work**  n=177 | **At school**  n=31 |
| --- | --- | --- |
| I fear judgment or stigma | 53 (29) | 8 (26) |
| I am concerned about being treated differently | 55 (29) | 9 (29) |
| I believe that the diagnosis is not relevant | 73 (41) | 14 (45) |
| I need formal or informal adjustments | 23 (13) | 5 (16) |
| I feel empowered by talking about it openly | 34 (18) | 5 (16) |
| I receive advice from family, friends or professionals | 14 (8) | 4 (13) |
| Other | 15 (8) | 3 (10) |
| No response | 3(1) | 1(3) |
| Results are presented as counts (n) and column percentages (%)  Additional facilitators and barriers mentioned by participants were: I feel protected by the law (n=1), I share it to guaranty the safety of my coworkers (n=1), I feel it is easier to manage my symptoms if others know (n=2), I am concerned it will affect my job security (n=2), People are not interested (n=2), I feel that when I share it I am admitting a weakness (n=1). | | |

Table S 5. Perceived impact of PCD on job security reported by adults and adolescents and retirement planning for adults. Responses from participants with primary ciliary dyskinesia registered in the Living with PCD study who completed the social functioning questionnaire

|  | **Adults and adolescents at work** |
| --- | --- |
| **Perceived impact of PCD in Job security** | **n=177** |
| Significantly | 23 (13) |
| Somewhat | 23 (13) |
| Neutral | 31 (17) |
| No, but I am concerned about it | 30 (17) |
| No, and I am nor concerned about it | 67 (38) |
| No response | 3 (2) |
| **Impact of PCD on retirement plans** | N = 39 |
| I had to retire earlier than planned | 28 (78) |
| I had to retire later than planned | 0 (0) |
| No impact | 11 (28) |
| Results are presented as counts (n) and column percentages (%) | |
